# In utero exposure to per – and polyfluoroalkyl substances (PFAS) associates with altered human infant T helper cell development

**DOI:** 10.1101/2024.11.18.24317489

**Authors:** Darline Castro Meléndez, Nathan Laniewski, Todd A. Jusko, Xing Qiu, B. Paige Lawrence, Zorimar Rivera-Núñez, Jessica Brunner, Meghan Best, Allison Macomber, Alena Leger, Kurunthachalam Kannan, Richard Kermit Miller, Emily S. Barrett, Thomas G. O’Connor, Kristin Scheible

## Abstract

**Background:** Environmental exposures to chemical toxicants during gestation and infancy can dysregulate multiple developmental processes, causing lifelong effects. There is compelling evidence of PFAS-associated immunotoxicity in adults and children. However, the effect of developmental PFAS exposure on infant T-cell immunity is unreported, and, if present, could be implicated in immune-related health outcomes.

**Objectives:** We seek to model longitudinal changes in CD4+ T-cell subpopulations from birth through 12 months and their association with in-utero PFAS exposure and postnatal CD4+ T-cell frequencies and functions.

**Methods:** Maternal-infant dyads were recruited as part of the UPSIDE-ECHO cohort during the first trimester between 2015 and 2019 in Rochester, New York; dyads were followed through the infant’s first birthday. Maternal PFAS concentrations (PFOS, PFOA, PFNA, and PFHXS) were quantified in serum during the second trimester using high-performance liquid chromatography and tandem mass spectrometry. Infant lymphocyte frequencies were assessed at birth, 6- and 12-months using mass cytometry and high-dimensional clustering methods. Linear mixed-effects models were employed to analyze the relationship between maternal PFAS concentrations and CD4+ T-cell subpopulations (n=200). All models included a PFAS and age interaction and were adjusted for parity, infant sex, and pre-pregnancy body mass index.

**Results:** In-utero PFAS exposure correlated with multiple CD4+ T-cell subpopulations in infants. The greatest effect sizes were seen in T-follicular helper (Tfh) and T-helper 2 (Th2) cells at 12 months. A log_2_-unit increase in PFOS was associated with lower Tfh [0.17% (95%CI: −0.30, −0.40)] and greater Th2 [0.27% (95%CI: 0.18, 0.35)] cell percentages at 12 months. Similar trends were observed for PFOA, PFNA, and PFHXS.

**Discussion:** Maternal PFAS exposures correlate with cell-specific changes in the infant T-cell compartment, including key CD4+ T-cell subpopulations that play central roles in coordinating well-regulated, protective immunity. Future studies into the role of PFAS-associated T-cell distribution and risk of adverse immune-related health outcomes in children are warranted.

## Introduction

The immune system development begins in the fetus during gestation and continues after birth. Different immune cell types arise from precursors, which can differentiate and develop in a regulated manner. ^1–3^ Environmental exposure to harmful agents such as chemical toxicants or deficits in nutrition during gestation and infancy can dysregulate multiple developmental processes influencing health over a lifetime.^4–7^ Among these harmful agents, exposure to per- and polyfluoroalkyl substances (PFAS) stands out due to their widespread prevalence and persistence in the environment, ^8^ and their well-documented associations with several adverse health outcomes across the lifespan, including impaired reproduction, growth, neurodevelopment and liver, kidney and immune dysfunction. ^9–14^ PFAS are a group of synthetic chemicals characterized by interconnected carbon and fluorine (C–F) atom chains. ^8^ The strong C–F bond makes PFAS resistant to environmental and physiological breakdown processes and allows them to accumulate and persist in the environment and human tissues. ^15^ In the U.S., over 97% of the population has detectable concentrations of PFAS in their blood^16^, and similar findings have been reported in studies from Spain,^17^ Germany,^18^ and China.^19^ In addition to blood, PFAS have been detected in human lungs, brain, bone, liver, and kidneys.^15^ PFAS have also been detected in the placenta, cord blood, and breast milk,^20–25^ which further emphasizes the potential for early life exposures.

One consistent observation is an inverse association between PFAS concentrations and antibody responses to vaccinations in adults and children. ^9,10,26–30^ It is unclear which immune system components are affected by PFAS to generate a net effect of lower humoral responses. The production of antibodies results from the collaboration between multiple types of immune cells.^31^ When the immune system is stimulated by vaccinations or infections, cells from the innate compartment, such as macrophages and neutrophils, trigger inflammatory processes that activate cells of the adaptive immune system (T and B-cells), which can then establish immune memory. Immune memory is the ability of cells to retain antigen-specific information from prior exposures to facilitate a faster and more targeted response upon subsequent encounters. Antibody production by B-cells is influenced by CD4+ T-cells, which consist of different specialized subpopulations tailored to the immune response needed for a specific pathogen or antigen.^32^ These include Th1, Th2, Th17, T follicular helper cells (Tfh), and T regulatory cells (Treg).^33^ Each CD4+ T-cell subset carries a unique set of functions that influence B-cells by modulating the strength, specificity, and durability of their produced antibodies. ^34^ For example, long-lived B-cells rely on Tfh cells for durable antibody production because Tfh cells promote germinal center formation in lymph nodes via IL-21 secretion.^35–40^ This interaction leads to B-cell proliferation, somatic hypermutation, and isotype class switching, ultimately resulting in high-affinity antibodies.^35,37^ Other CD4+ T-cell subpopulations modulate antibody class switching by fine-tuning the context of the immune response required.^41–44^ How early life exposures to PFAS affects these cell types is crucial for discovering how PFAS ultimately leads to changes in immune responses. This longitudinal study aims to identify alterations in CD4+ T-cells related to developmental PFAS exposure as a first step in understanding pathways implicated in PFAS-related immune dysfunction.

## Methods

### Study design

Between December 2015 and April 2019, first-trimester pregnant women were recruited from outpatient clinics affiliated with the University of Rochester and enrolled in the Understanding Pregnancy Signals and Infant Development and Environmental Influences on Child Health Outcomes (UPSIDE-ECHO) study after informed consent. UPSIDE-ECHO is an ongoing prospective birth cohort designed to study prenatal exposures in relation to perinatal and child health outcomes in the Rochester, NY, area^45^ that is part of the NIH ECHO program.^46^ Eligibility criteria included maternal age 18 years or older, singleton pregnancy, no known substance abuse problems or history of psychotic illness, and the ability to communicate in English. Participants with major endocrine disorders (e.g., polycystic ovary syndrome) and high-risk pregnancies at baseline were not enrolled; additionally, infants born prior to 37 weeks of gestation or with congenital anomalies or significant health problems were excluded (n=9). Study visits, including physical exams, biospecimen collection, and questionnaire completion, were completed at each trimester and at birth, 6 and 12 months postnatally. This study protocol was approved by the University of Rochester School of Medicine and Dentistry Institutional Review Board (#58456).

### Maternal and infant blood collection

During the second trimester of pregnancy (21 ± SD:1.8) gestational weeks), maternal serum from blood collected into EDTA tubes was frozen at −80°C until the time of analysis. For infant blood samples, cord blood was collected by venipuncture into sodium heparin tubes immediately after birth. Infant blood was collected into sodium heparin tubes at 6 and 12 months. Peripheral blood mononuclear cells (PBMC) were isolated using a Ficoll-Paque density gradient (GE Healthcare – cytiva) and immediately cryopreserved in freezing media consisting of 10% heat-inactivated Fetal Bovine Serum (FBS)(Seradigm) and 10% DMSO (Amresco) at a cell concentration ranging from one to five million cells per mL.

### PFAS analysis

Maternal serum isolated from the second trimester of pregnancy was sent to Wadsworth Center’s Human Health Exposure Analysis Resource (HHEAR) Laboratory Hub for PFAS measurement. PFAS were measured using high-performance liquid chromatography (HPLC) and tandem mass spectrometry (MS/MS) detection and quantification, as described in Honda et al. (2018).^47^ Fourteen PFAS were measured: NETFOSAA, NMFOSAA, PFBS, PFDA, PFDODA, PFHPA, PFHXA, PFHXS, PFNA, PFOA, PFOS, PFOSA, PFPEA, PFUNDA. The limit of detection (LOD) for individual PFAS varied from 0.02 to 0.03 ng/mL. To reduce exposure misclassification, four PFAS with 100% detection rate, namely PFOA, PFOS, PFNA, and PFHXS were included in the present analysis. Several quality control samples namely certificated standard reference materials (NIST SRM 1957 and SRM 1958), HHEAR QC pools, procedural blanks and matrix spikes were included in the analysis. The coefficient of variation in concentrations of four PFAS in SRMs, QC pools and matrix spikes between batches was <10%.

### Infant T-cell phenotyping

Thawing, stimulation, and staining of PBMCs for mass cytometry were performed as detailed previously Scheible et al. (2012) and McDavid et al. (2022).^48,49^ To control for batch effect and variability, all timepoints (birth, 6, and 12 months) from individual infant subjects were phenotyped in experimental batches that included 20 individual samples maximum. Each experimental batch also included one to two healthy adult PBMC controls that were carried across multiple batches. Cryopreserved sample vials were thawed at 37°C in a water bath and washed twice with 10mL of RPMI 1640 (Corning) supplemented with L-glutamine, 10% heat-inactivated Fetal Bovine Serum (FBS) (R10), and 25U/uL Universal Nuclease (R10+n). Cord blood samples (CBMC) were treated with 3 mL 1x BD Pharm Lyse (BD Biosciences) supplemented with 25U/uL Universal Nuclease for 15 minutes to lyse red blood cells. Washed cells were rested overnight at 37°C in a 5% CO_2_ incubator. After overnight rest, samples were counted on a dual-fluorescence cell counter (Cellometer K2; Nexcelom/Revvity); samples less than 70% viable were excluded from the assay (5.9% of samples at birth, 14.6% at 6 months and 12.7% at 12 months). Cells were resuspended in R10 at a concentration of 30 million-cells/mL, and 3 million cells were stimulated for 2 hours with 2ug/mL of *Staphylococcus aureus*, Enterotoxin Type B (SEB). GolgiPlug (BD Biosciences) and monensin (Sigma; 10 mM stock in EtOH) were added at 2 hours and further stimulated for 8 hours at 37°C. Stimulated cells were washed with stain buffer (PBS + 2%FBS + 25U/uL Universal Nuclease) (SBn) and stained for 30 minutes at room temperature (RT) with unique combinations of metal-conjugated anti-human CD45 antibodies (Supplemental Table 1: Parameter Type, Barcodes) in a ‘6-choose-3’ or ‘7-choose-3’ live-cell barcoding scheme. ^50–52^ Barcoded samples were washed, resuspended in SBn, and then transferred into a single 15mL culture tube. The pooled sample was filtered, centrifuged, and stained for 30 minutes at RT with a cocktail of metal-conjugated anti-human antibodies for surface molecules (Supplemental Table 1: Parameter Type, Surface). Next, cells were washed with SBn then stained with Cell-ID Cisplatin-194Pt (Fluidigm/Standard BioTools) at 500nM/mL concentration for 5 minutes at RT. Cells were washed with SBn and fixed/permeabilized for 30 minutes at RT with Foxp3 Fixation-Permeabilization buffer (eBioscience) followed by a wash with Permeabilization Buffer (Perm-B) (eBioscience). Cells were intracellularly stained for 30 minutes at RT with Perm-B containing a cocktail of metal-conjugated anti-human antibodies (Supplemental Table 1: Parameter Type, Intracellular). Cells were washed, then fixed with 1 mL of 1.6% formaldehyde (Pierce) in PBS containing 500 nM of Cell-ID Intercalator-103Rh (Fluidigm/Standard BioTools) and incubated overnight at 4°C. Fixed cells were washed with PBS, counted, and resuspended at a concentration of 13.75 million-cells/mL in filtered FBS with 10% DMSO (Amresco). Cells were aliquoted in Protein Lo-Bind tubes (Eppendorf) (2.75 million cells in 200ul) and stored at −80°C. Following aliquoting and freezing, individual aliquots were prepared for acquisition on a CyTOF 2 or Helios mass cytometer (Fluidigm/Standard BioTools) by washing twice with 2 mL ultrapure water and resuspending in 3 mL ultrapure water containing 0.1x EQ Four Element Calibration Beads (Fluidigm/Standard BioTools). Aliquots were acquired using a Super Sampler (Victorian Airships).

#### Mass cytometry clustering pipeline

After the acquisition, source data was normalized using the Normalization Passport in CyTOF Software with default settings (Fluidigm, 7.0.5189). Normalized data was analyzed using a custom R-based workflow (github.com/nlaniewski/SOMnambulate). Briefly, unique live-cell barcoded pools were debarcoded to generate sample-specific *.fcs* files. FlowSOM^53^ was used to generate clusters representing major PBMC immune cell populations (Supplemental Table 1: FlowSOM dimension, PBMC). Based on per-cluster feature expression, annotations were defined. Selecting for only CD3+, CD4+, and CD8+ clusters representing T-cells, a second FlowSOM was generated using an expanded set of both surface and intracellular markers to define T-cell-specific sub-clustering results (Supplemental Table 1: FlowSOM dimension, T-cells). As with the PBMC clusters, per-cluster feature expressions were examined to define functional annotations. To aid in the interpretation of FlowSOM clusters, the nodes that comprise the self-organizing map (SOM) were visualized using Uniform Manifold Approximation and Projection (UMAP).^54^ Default parameters were used to embed the nodes into two dimensions, using the same markers as those used to generate the respective SOM. Nodes (circles) were individually sized to represent the mean proportion of assigned cells and colored by their assigned cluster annotation. For each FlowSOM result (PBMC; T-cell), all cells were mapped on a per-sample basis, and the proportion of cells attributed to each cluster was calculated. The proportions used in subsequent visualizations and statistical models represent the percent of CD4+ T cells.

### Measurement of covariates and selection of potential confounding variables

After study enrollment in the first trimester, trained staff administered a questionnaire to obtain information on lifestyle, living environment, past pregnancies, medical conditions, medications used before, during, and after pregnancy, and sociodemographic information. Sociodemographic information, including self-reported maternal race and ethnicity, was collected at baseline, recognizing that these social constructs are associated with environmental exposures and maternal and child health outcomes.^55,56^ Infant birth weight and sex at birth were obtained from electronic health records. Confounding variables were selected using a directed acyclic graph (DAG) informed by previous reports demonstrating an association with PFAS and T-cells (Supplemental Figure 1).^10,57^ These were the categorical variables parity, infant sex, mode of delivery, maternal race, maternal education, and Medicaid during pregnancy, and the continuous variables maternal age, pre-pregnancy body mass index (BMI), annual income, and infant birth weight. The minimally sufficient set of variables from the DAG were categorical parity (0, 1, or 2+), infant sex at birth, and continuous pre-pregnancy BMI.

### Statistical analysis

Descriptive characteristics for the entire sample are presented in Table 1. Specifically, frequencies and percentages for categorical variables, as well as means with standard deviations for continuous variables are reported in this table. Wilcoxon rank-sum test was used for comparing mean differences between two groups for a continuous variable, Wilcoxon signed-rank test was used to assess mean differences between two time points. The association between two continuous variables was evaluated using Spearman’s rank correlation test.

**Table 1:**
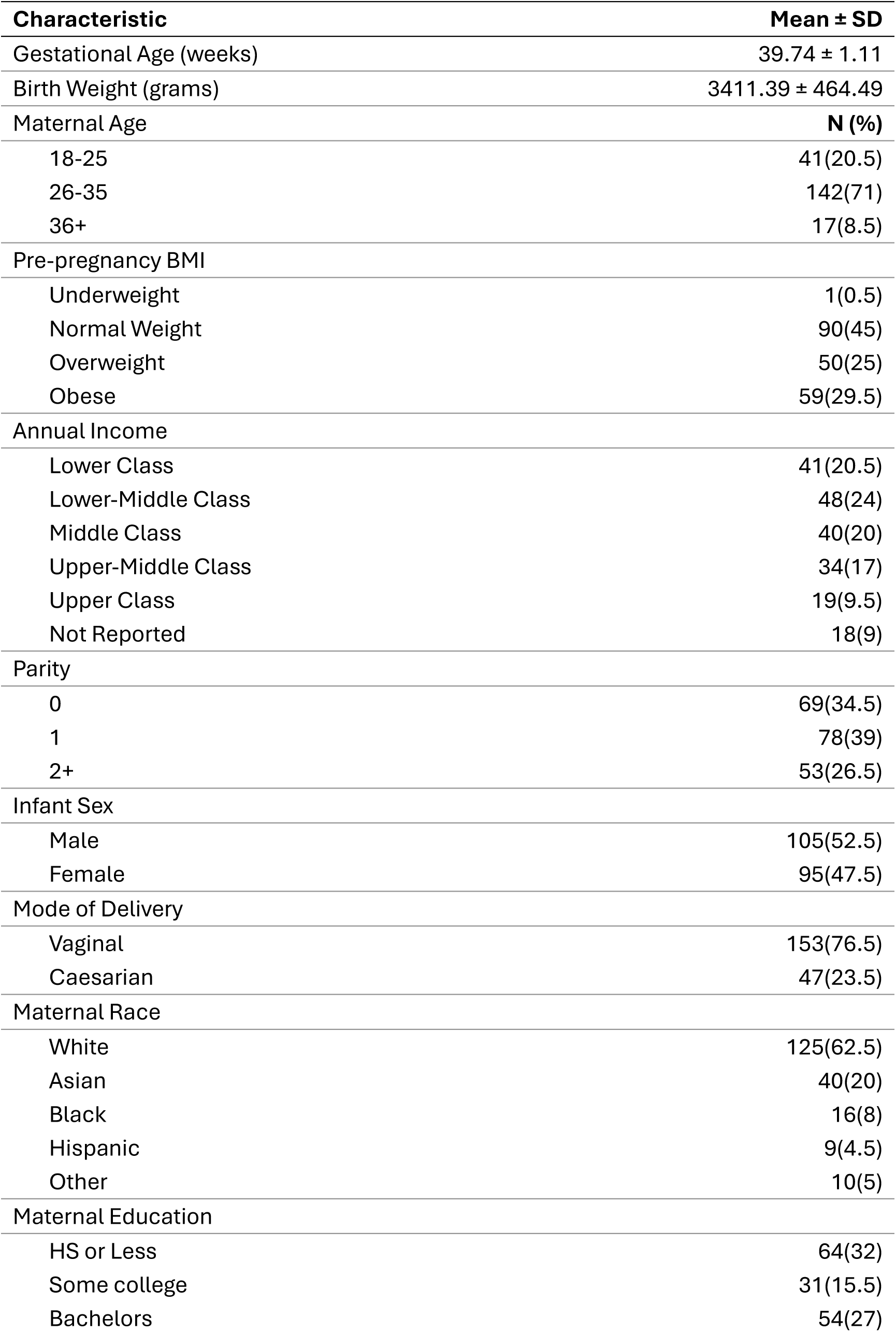
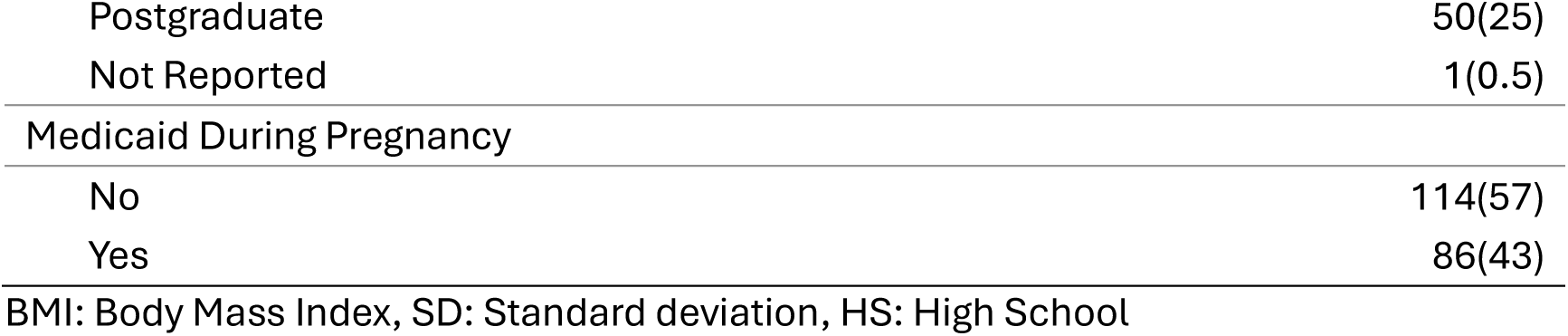
Infant and maternal characteristics. (n=200)

| **Characteristic** | **Mean ± SD** |
| --- | --- |
| Gestational Age (weeks) | 39.74 ± 1.11 |
| Birth Weight (grams) | 3411.39 ± 464.49 |
| Maternal Age | **N (%)** |
| 18-25 | 41(20.5) |
| 26-35 | 142(71) |
| 36+ | 17(8.5) |
| Pre-pregnancy BMI |  |
| Underweight | 1(0.5) |
| Normal Weight | 90(45) |
| Overweight | 50(25) |
| Obese | 59(29.5) |
| Annual Income |  |
| Lower Class | 41(20.5) |
| Lower-Middle Class | 48(24) |
| Middle Class | 40(20) |
| Upper-Middle Class | 34(17) |
| Upper Class | 19(9.5) |
| Not Reported | 18(9) |
| Parity |  |
| 0 | 69(34.5) |
| 1 | 78(39) |
| 2+ | 53(26.5) |
| Infant Sex |  |
| Male | 105(52.5) |
| Female | 95(47.5) |
| Mode of Delivery |  |
| Vaginal | 153(76.5) |
| Caesarian | 47(23.5) |
| Maternal Race |  |
| White | 125(62.5) |
| Asian | 40(20) |
| Black | 16(8) |
| Hispanic | 9(4.5) |
| Other | 10(5) |
| Maternal Education |  |
| HS or Less | 64(32) |
| Some college | 31(15.5) |
| Bachelors | 54(27) |
| Postgraduate | 50(25) |
| Not Reported | 1(0.5) |
| Medicaid During Pregnancy |  |
| No | 114(57) |
| Yes | 86(43) |

BMI: Body Mass Index, SD: Standard deviation, HS: High School

Owning to the longitudinal assessment of T-cell outcomes at multiple visits, two separate but related Linear Mixed-Effect regression (LMER) modeling strategies were employed to associate T-cell outcomes with individual PFAS concentrations, infant age, and their interactions. In both models, infant age is considered as a categorical variable with three levels (birth, 6 and 12 months), of which birth is considered as the baseline. In the first set of LMER models, a log-transformation designed to reduce the skewness is applied to the PFAS concentration levels: *x*_*ij*_ = log_2_(Conc_*ij*_ + 1), where Conc_*ij*_ represents the original PFAS concentration level for the *i*th subject at the *j*th visit. In the second set of LMER models, PFAS concentration levels were dichotomized as “above median” or “below median”, permitting the estimation of differences in T-cell outcomes over time as discrete categories according to the categorical interactions between infant age and binary PFAS concentration levels. We call the second set of models as differences-over-time models.

For both approaches, a model is called a *crude model* if it only includes a PFAS level, infant age, and their interactions. It is called an *adjusted model*, if it also includes the following additional fixed effects: parity (0, 1, or 2+), infant sex, and pre-pregnancy BMI (continuous). A subject-specific random intercept was included in all models to account for serial correlation in the repeated T-cell measures. Unknown parameters in these models were estimated by the Restricted maximum likelihood (REML) criterion. Regression t-test implemented in R package *lmerTest*^58^ was used to assess the statistical significance of a fixed effect of interest; *F*-tests as implemented in function linearHypothesis() in R package *car* ^59^ was used to test specific linear contrasts between infant ages. Results for the difference-over-time models were presented as differences in marginal mean percent with corresponding 95% confidence intervals, and pairwise comparisons were performed to contrast mean percentages between below and above median PFAS. Model assumptions, including normality, linearity, and homoscedasticity of the residuals, were visually evaluated by inspection of diagnostic plots. A p-value of less than 0.05 was considered statistically significant for individual hypothesis tests. For analyses involving multiple hypotheses, the Benjamini–Hochberg procedure was employed to control the false discovery rate (FDR) at a level of 0.05. All statistical analyses were conducted using R Foundation for Statistical Computing version 4.3.3.^60^

## Results

### Study Population

The analysis included 200 full-term infants (Supplemental Figure 2). Participants were selected from the larger UPSIDE-ECHO cohort if they had second-trimester maternal PFAS measured and infant T-cell phenotyping from at least one timepoint (birth, 6, or 12 months of age). Cohort characteristics are shown in Table 1. In summary, 71% of participants were between 26-35 years old at enrollment; 62.5% of the cohort self-identified as White, 20% as Asian, 8% as Black, 9% as Hispanic, and 10% as other. 52% reported having at least a bachelor’s degree, and 43% were enrolled in Medicaid during pregnancy. Parity was categorized based on pregnancies preceding the current pregnancy (34.5% nulliparous, 39% primiparous, or 26.5% multiparous). The mean gestational age at birth was 39.74 ± SD:1.11 weeks, 76.5% of infants were born vaginally, and 52.5% were assigned male sex at birth. Among the 200 mother-infant pairs who contributed at least one observation to the current study, compared to birth, those with 6- or 12-month data tended to be older at the time of the child’s birth, higher income and education, and more likely to report White race (Supplemental Table 2).

### PFAS exposure

Table 2 shows the mean concentration and distribution of four PFAS (PFOS, PFOA, PFNA, and PFHXS) that were detected in 100% of the second-trimester maternal serum samples and, therefore, included in subsequent analysis. The concentrations and distribution of the ten additional PFAS measured are in Supplemental Table 3. PFOS was present at the highest mean concentration (2.87±2.13 ng/mL), followed by PFHXS (1.96±0.92 ng/mL), PFOA (0.68±0.51 ng/mL), and the lowest concentration being PFNA (0.33±0.36 ng/mL). Relative to the NHANES general population concentrations, PFOS, PFOA, and PFNA mean concentrations were lower. ^61^ PFOA and PFNA were the most strongly correlated (Rho=0.64, p-value= 0.0001), followed by PFOS and PFNA (Rho=0.58, p-value= 0.0002) and PFOS and PFOA (Rho = 0.53, p-value= 0.0005). PFHXS showed a moderate correlation with PFOS (Rho= 0.38, p-value= 0.021) and PFOA (Rho= 0.30, p-value =0.049); PFNA and PFHXS were the most weakly correlated (Rho= 0.23, p-value = 0.113).

**Table 2:**
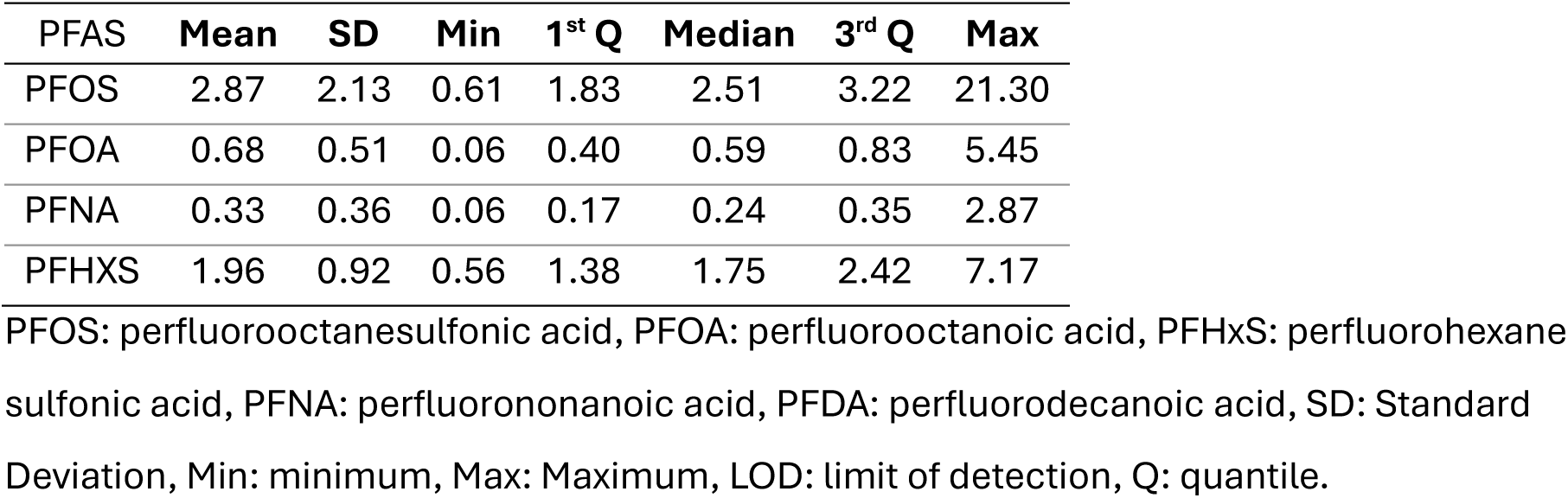
Distribution of perfluoroalkyl substances (PFAS) in maternal plasma (ng/mL).

| PFAS | Mean | SD | Min | 1<sup>st</sup> Q | Median | 3<sup>rd</sup> Q | Max |
| --- | --- | --- | --- | --- | --- | --- | --- |
| PFOS | 2.87 | 2.13 | 0.61 | 1.83 | 2.51 | 3.22 | 21.30 |
| PFOA | 0.68 | 0.51 | 0.06 | 0.40 | 0.59 | 0.83 | 5.45 |
| PFNA | 0.33 | 0.36 | 0.06 | 0.17 | 0.24 | 0.35 | 2.87 |
| PFHXS | 1.96 | 0.92 | 0.56 | 1.38 | 1.75 | 2.42 | 7.17 |

PFOS: perfluorooctanesulfonic acid, PFOA: perfluorooctanoic acid, PFHxS: perfluorohexane sulfonic acid, PFNA: perfluorononanoic acid, PFDA: perfluorodecanoic acid, SD: Standard Deviation, Min: minimum, Max: Maximum, LOD: limit of detection, Q: quantile.

Determinants of exposure for the UPSIDE-ECHO population can be found in Supplemental Table 4. Overall, the concentrations of all PFAS were lower, with higher maternal parity. Maternal PFAS concentrations in serum were lower when the fetal sex was female. Generally, mean PFAS concentrations were higher in white mothers and mothers with higher income and education, and mean PFAS concentrations were lower in older mothers and mothers with higher pre-pregnancy BMI. Participants with Medicaid during pregnancy had lower concentrations of PFAS.

### Infant T-cell clustering and phenotyping

A 36-marker mass cytometry panel (Supplemental Table 1) was used to deeply immunophenotype infant immune cell populations over the first year of life. FlowSOM clustering algorithm was used to identify and group immune populations (Figure 1A). PBMC populations were identified as follows: γδ T-cells were identified as CD3+ TCRγδ+ cells, cytotoxic T-cells as CD3+ CD8+, helper T-cells as CD3+ CD4+, B-cells as CD19+, NK cells as CD56+, and monocytes as CD14+ cells. UMAP visualization of infant leukocytes revealed separation between the major immune populations, including T-cells, B-cells, NK cells, and monocytes (Figure 1B).

**Figure 1:**
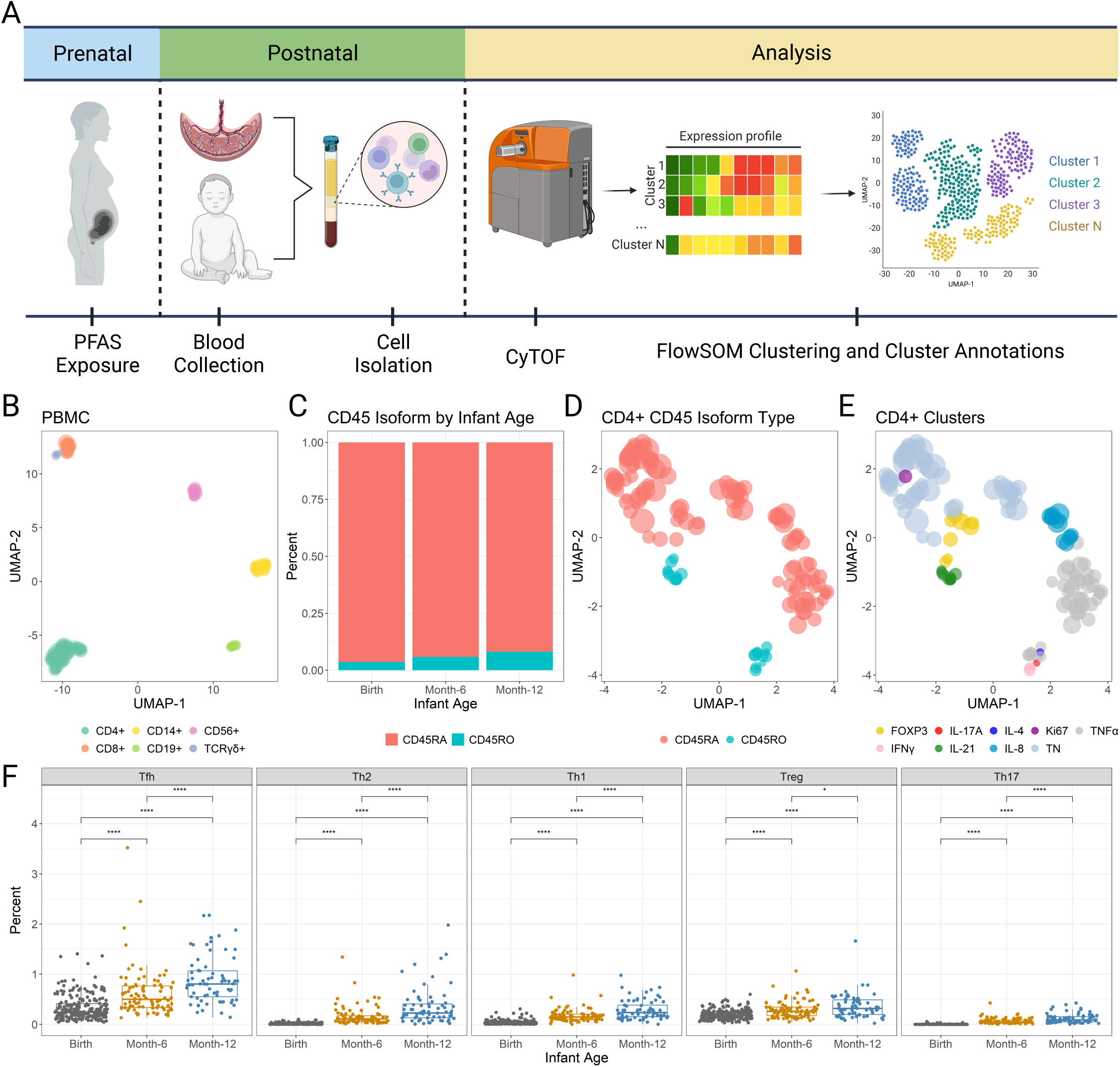
Identification of CD4+ T-cell subpopulations in infants and their age-associated frequencies. (A) Study design and T-cell analysis pipeline. (B) Uniform Manifold Approximation and Projection (UMAP) representation of FlowSOM nodes colored by major peripheral blood mononuclear cells (PBMC): CD4+, CD8+, CD14+, CD19+, CD56+, TCRγδ; (C) Stacked bar graph denoting the proportion of CD45RA+ and CD45RO+ CD4+ T-cell subpopulations at birth, 6 and 12 months of age. (D-E) UMAP representation of FlowSOM nodes colored by expression of (D) CD45 isoform type (CD45RA+ or CD45RO+) within CD4+ T-cell FlowSOM nodes, and (E) CD4+ T-cell sub-cluster annotations based on the following marker expression: Treg (FOXP3+CD127-), Th1(IFNγ+), Th17 (IL-17A+), Tfh (IL-21+), Th2 (IL-4+), Naïve-cytokine-negative (TN). Circle size on UMAPs represents the relative mean count of total cells assigned to each FlowSOM node. (F) Frequency of CD4+ T-cells in cord blood at birth and PBMC at six and twelve months. Each dot on the boxplots denotes data from an individual infant at the indicated age; scales for boxplots are fixed.

To test our specific hypothesis regarding the role of PFAS in T-cell development, we narrowed analyses to CD4+ T-cells. CD4+ T-cells from the first level of clustering (i.e., PBMC) were then selected for a second level of FlowSOM clustering using an expanded set of phenotypic and functional markers to identify unique CD4+ T-cell sub-clusters. This sub-clustering analysis identified 29 unique CD4+ T-cell sub-clusters (Supplemental Figure 3A-B). During the first year of life, naïve (CD45RA+) comprised most of the CD4+ T-cell compartment, while T-cells carrying markers of immunological memory (CD45RO+, CD45RA-) were a small fraction at birth and increased over time (Figure 1C-D). Within the memory (CD45RO+) compartment, canonical CD4+ T-cell subpopulations Th1 (IFNg+), Th2 (IL-4+), Th17 (IL-17A+), Tregs (FOXP3+ CD127-) and Tfh cells (IL-21+) were identified based on the expression of surface and intracellular markers that distinguish functional properties of these subpopulations (Figure 1E, Supplemental Figure 3B-C). The percentages of these CD4+ T-cell subpopulations increased over time (Figure 1F, Supplemental Table 5).

### Maternal PFAS exposure associates with infant CD4+ T-cell frequencies in an age-dependent manner

Figure 2 shows the age-specific adjusted association for PFOS, PFOA, PFNA, and PFHXS and each CD4+ T-cell. The unadjusted and adjusted age-specific estimates for this model can be found in Supplemental Table 6. Overall, the associations between PFAS and CD4+ T-cells were stronger at 6- and 12-months of age versus birth; with statistical evidence of effect measure modification by age for Tfh, Th2, and Th1 cells, but not for Tregs or Th17 cells (Supplemental Table 7 and Supplemental Table 8). These associations were consistent for crude and adjusted models.

**Figure 2:**
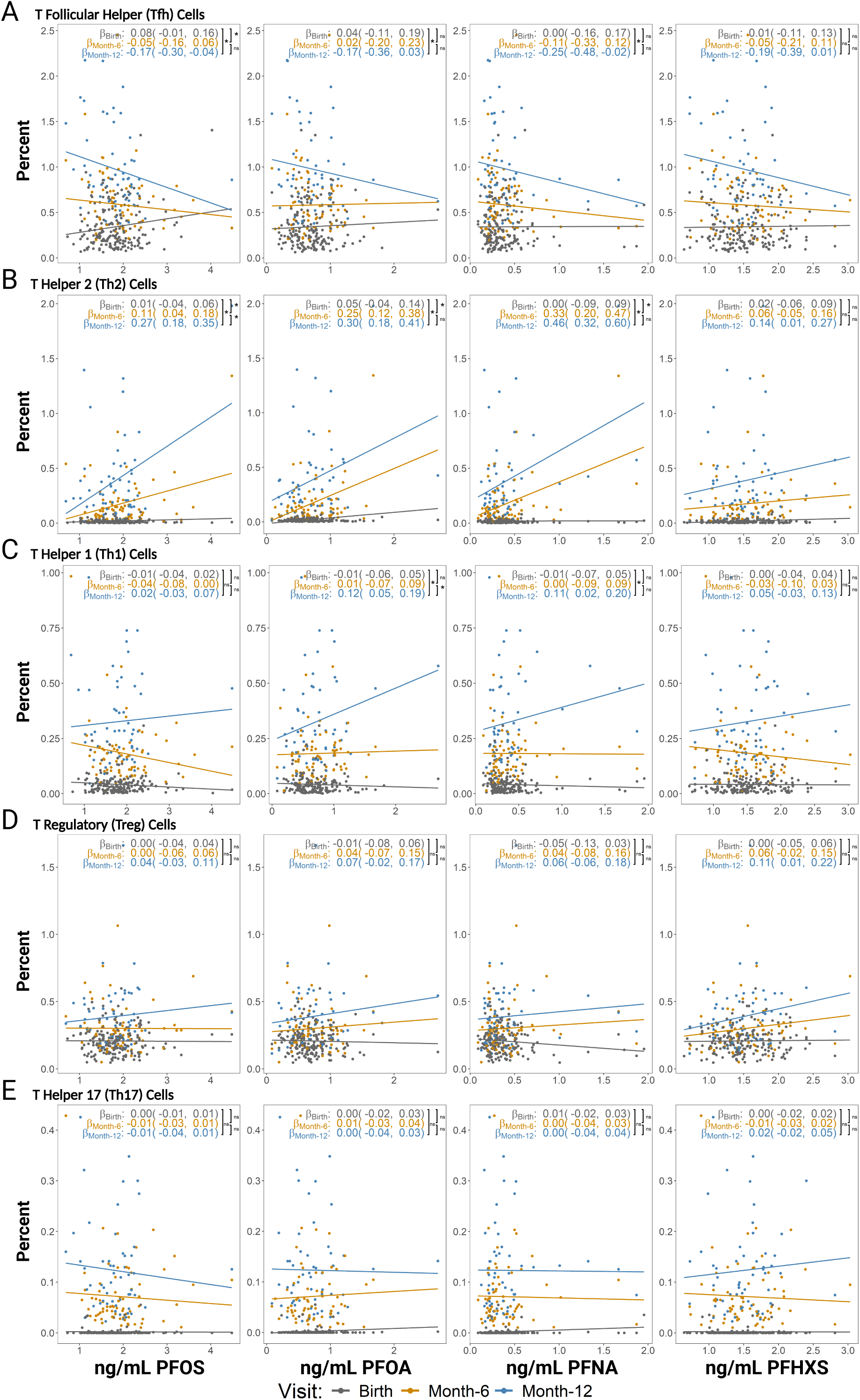
Maternal PFAS exposure associates with infant CD4+ T-cell frequencies in an age-dependent manner. The plots illustrate the regression lines for the age-specific average marginal effects of the association between continuous maternal PFAS (ng/mL) exposure and infant CD4+ T-cell percentage. Rows represent individual CD4+ T-cells: (A) T follicular helper cells, (B) T helper 2 cells, (C) T helper 1 cells, (D) T regulatory cells, and (E) T helper 17 cells. Columns represent individual PFAS; PFOS, PFOA, PFNA, and PFHXS were log_2_ transformed. The colors represent the infant age: birth (grey), 6 months (orange), and 12 months (blue). Each dot represents an individual infant at the indicated age. Age-specific average marginal effects (β) were adjusted for the categorical covariates parity, infant sex, and continuous pre-pregnancy body mass index. Parenthesis next to each age-specific β contains the 95% confidence interval for the estimate. Pairwise comparisons between the age-specific βs are represented by the brackets. Asterisks denote statistically significant (p-value < 0.05), and ‘ns’ indicates non-statistically significant differences between age-specific βs. Axis are independent for each CD4+ T-cell subpopulation and each PFAS. Supplemental Tables 6, 7, and 8 contain corresponding numeric data.

Tfh cells were inversely associated with maternal PFAS; each doubling of PFOS was associated with a 0.17 (95%CI of β: −0.30, −0.04) lower Tfh percentage measured at 12 months of age, a 0.05 (95%CI of β: −0.16, 0.06) lower Tfh percentage at 6 months, and a 0.08 (95%CI of β: −0.01, 0.16) greater Tfh percentage at birth (p-value for age interaction=0.002). Similar trends were observed between Tfh percentages and PFOA, PFNA and PFHXS (Figure 2A).

In contrast, Th2, Th1, and Treg cells were positively associated with maternal PFAS exposure. For Th2 cells, each doubling of PFOA was associated with a 0.30 (95%CI of β: 0.18, 0.41) greater Th2 percentage measured at 12 months of age, a 0.25 (95%CI of β: 0.12, 0.38) greater Th2 percentage at 6 months, and a 0.05 (95%CI of β: −0.04, 0.14) greater Th2 percentage at birth (p-value for age interaction = 0.000). Similar trends were observed between Th2 percentages and PFOS, PFNA, and PFHXS (Figure 2B). For Th1 cells, each doubling of PFNA was associated with a 0.11 (95%CI of β: 0.02, 0.20) greater Th1 percentage measured at 12 months of age and no association at 6 months of age or birth (p-value for age interaction = 0.054). Similar trends were observed between Th1 percentages and PFOS, PFOA, and PFHXS (Figure 2C).

For Tregs, each doubling of PFHXS was associated with a 0.11 (95%CI of β: 0.01, 0.22) greater Treg percentage measured at 12 months of age, a 0.06 (95%CI of β: −0.02, 0.15) greater Treg percentage at 6 months, and no association at birth (p-value for age interaction = 0.129). Similar trends were observed between Treg percentages and PFOS, PFOA, and PFNA (Figure 2D). No associations were observed between Th17 percentages and the doubling of PFOS, PFOA, PFNA, and PFHXS (Figure 2E).

### Maternal PFAS exposure predicts CD4+ T-cell trajectories in infants

Overall, independently of PFAS exposure group, the trajectory of each CD4+ T-cell subset showed increasing levels over time (Figure 3). Further, PFAS-dependent effects were more pronounced at later ages, especially 12 months of age. This was the case for Tfh, Th2, Th1, and Treg cells, but not for Th17 cells. Unadjusted and adjusted age-specific marginal means and pairwise comparisons for below and above median PFAS can be found in Supplemental Tables 9 and 10, respectively. For Tfh cells and PFOS exposure, the adjusted negative association was strongest at 12 months of age, with a mean value for above median PFOS of 0.86 ± SD: 0.05 (95%CI:0.76, 0.96) versus a mean value for below median PFOS of 1.13 ± SD: 0.06 (95%CI:1.02, 1.24) (p-value for difference=0.002); similar trends were observed for PFOA and PFNA, and no differences were observed for PFHXS (Figure 3A). For Th2 cells and PFOA exposure, the adjusted positive association was present at 6 months but stronger at 12 months of age. At 12 months we observed a mean value for Th2 cells for above median PFOA of 0.44 ± SD: 0.03 (95%CI:0.37, 0.50) versus a mean value for below median PFOA of 0.32 ± SD: 0.04 (95%CI: 0.25, 0.39) (p-value for difference = 0.002); similar trends were observed for PFOS, PFNA, and PFHXS (Figure 3B). For Th1 cells and PFNA exposure, the adjusted positive association was observed at 12 months of age, with a mean value for above the median PFNA of 0.37 ± SD:0.02 (95%CI: 0.32, 0.41) versus a mean value for below median PFNA of 0.30 ± SD: 0.02 (95%CI: 0.25, 0.34) (p-value for difference = 0.019); similar trends were observed for PFOS, PFOA, and PFHXS (Figure 3C). For Treg cells and PFHXS exposure, the adjusted positive association was present at 6 months but stronger at 12 months of age; at 12 months we observed a Treg mean value for above the median PFHXS of 0.45 ± SD: 0.03 (95%CI: 0.40, 0.50) versus a mean value for below the median PFHXS of 0.32 ± SD: 0.03 (95%CI: 0.26, 0.38) (p-value for difference = 0.002); similar trends for 12 months but not 6 months were observed for PFOS, PFOA, and PFNA (Figure 3D). No differences between the mean value of Th17 cells for above versus below median PFOS, PFOA, PFNA, and PFHXS were observed (Figure 3E).

**Figure 3:**
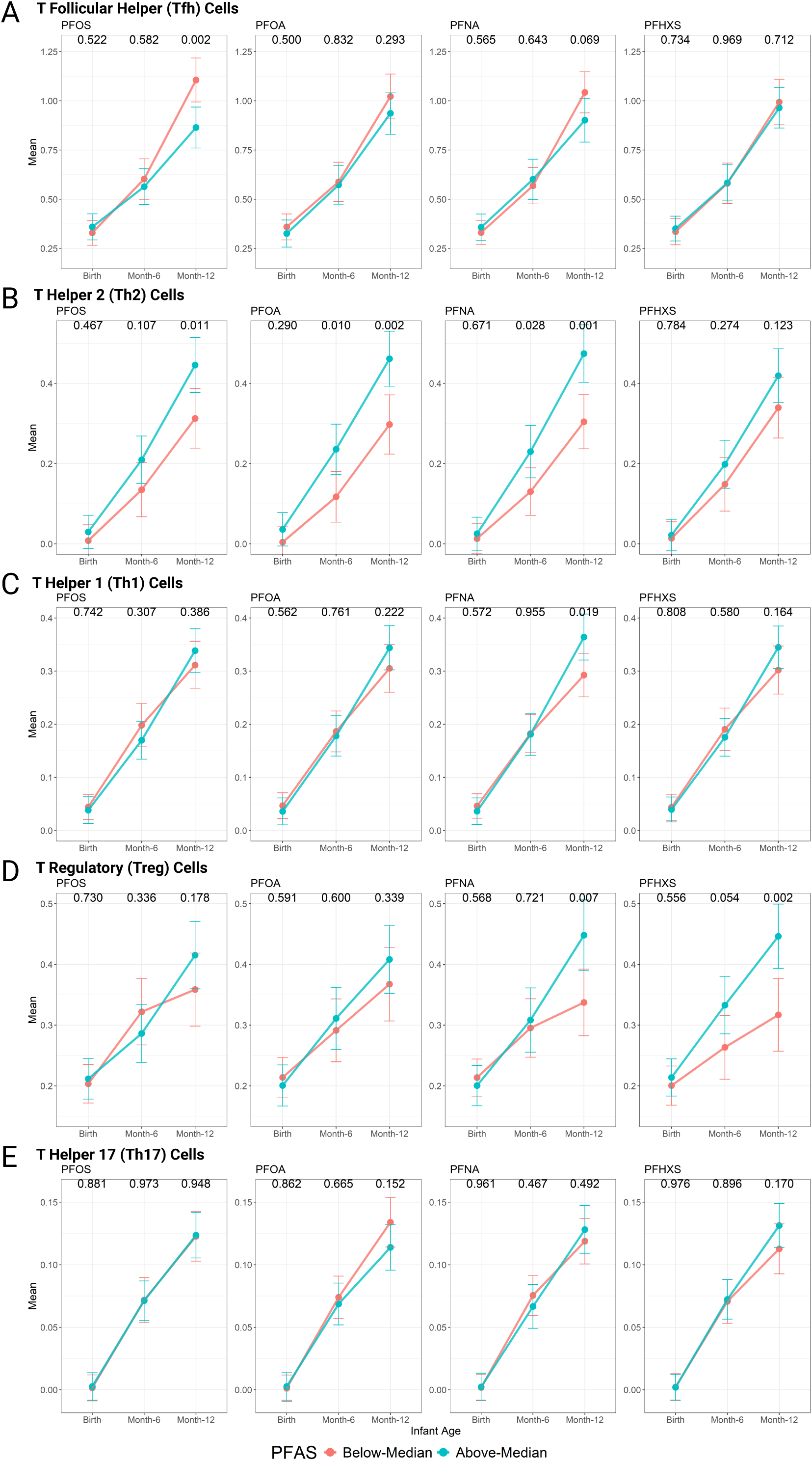
Maternal PFAS exposure predicts CD4+ T-cell trajectories in infants. The plots illustrate differences-over-time between infant CD4+ T-cell subpopulations by a categorical PFAS value (below versus above median value). Rows represent individual CD4+ T-cells: (A) T follicular helper cells, (B) T helper 2 cells, (C) T helper 1 cells, (D) T regulatory cells, and (E) T helper 17 cells. Columns represent individual PFAS (PFOS, PFOA, PFNA, and PFHXS). The colors represent the categorical PFAS value: below the median (pink) and above the median (blue). Dots denote the predicted marginal mean of each CD4+ T-cell subpopulation at the indicated age. Marginal means were adjusted for the categorical covariates parity, infant sex, and continuous pre-pregnancy body mass index. Error bars represent the standard error of the marginal means. P-values denote the statistical significance between marginal means contrasting below vs. above the median PFAS; the contrast was considered statistically significant if the p-value < 0.05. The y-axis is independent for each CD4+ T-cell subpopulation. Supplemental Tables 9, 10, and 11 contain corresponding numeric data.

## Discussion

In this prospective cohort study, we investigated the association between maternal PFAS exposure CD4+ T-cell subpopulations in infants over the first year of life. The potential of PFAS to affect the immune system has been widely reported in population studies across the world, with consistently observed reductions in antibody responses to common childhood vaccinations.^9,12^ Yet, the specific immune cell components altered by PFAS exposure remain unknown. We found that increased concentrations of maternal PFAS were associated with a reduction in the percentage of Tfh cells, increased Th2, Th1, and Treg populations, and no differences in Th17 cells. Moreover, each association was stronger from birth to 12 months of age. These findings indicate that the potential effects of prenatal PFAS exposure may become more pronounced as the infant grows older. This could be attributed to ongoing T-cell development in the first year of life, and that certain T-cell subpopulations may not emerge until the infant is exposed to specific stimuli. As time passes and T-cell subpopulations continue to develop, the impact of PFAS exposure on the cells becomes more apparent.

The immune system protects against pathogens through a coordinated network of cells, tissues, and organs. CD4+ T-cells orchestrate the immune response, making it specific to the type of stimuli received and aiding in the generation and maintenance of immune memory. Particularly, Tfh cells help direct B-cells to produce long-lasting, high-affinity antibodies essential for immune memory.^35^ This study revealed a negative association between the percentage of Tfh cells and maternal PFAS exposure during pregnancy. A reduction in Tfh cells in infants is consistent with the observed reductions in antibodies to childhood vaccinations.^27,28,62–66^ This provides potential mechanistic insights into this observation because Tfh cells are essential for mature and durable antibody responses.^35,39,40,67^

Given the complexity of antibody responses, it is unlikely Tfh cells are the only immune cell type altered by PFAS exposure. For example, we also observed that Tregs were enhanced with increased maternal PFAS concentrations. Tregs are recognized for their immunosuppressive functions and their capacity to inhibit the activation and proliferation of effector T-cells.^68,69^ Studies have demonstrated that Tregs can reduce immune responses to vaccinations by suppressing effector responses.^70,71^ An increase in Treg cells could suppress T-cell or B-cell responses, thereby attenuating antibody production. More generally, an enrichment in Treg cells could support an immunosuppressed environment, dampening the overall ability to fight infections. Therefore, an increased proportion or function of Tregs could also contribute to reduced antibody titers that have been observed in association with PFAS exposure.

In addition to Tfh and Tregs, our study found a positive association between the percent of Th1 and Th2 cells with maternal PFAS concentrations. While Th1 and Th2 cells play important defense roles during infections, excessive Th1 and Th2 activity can be involved in the immunopathogenesis of several chronic health conditions, including allergy and autoimmunity.^72–77^ Th1 cells are among the predominant T-cell subpopulations in systemic autoimmune disorders, such as rheumatoid arthritis^78^, as well as organ-specific autoimmunity, such as Type 1 diabetes mellitus^79^, and other debilitating inflammatory disorders such as Crohn’s disease.^80^

On the other hand, Th2 cells have been associated with the development of atopic and allergic diseases such as atopic dermatitis, eczema, chronic rhinosinusitis, and asthma.^77,81,82^ While studies have associated PFAS exposure with allergic or autoimmune outcomes in children, the findings are inconsistent, with some showing no association.^83–94^ Several studies examining the association between PFAS exposure and allergic outcomes in infants have measured IgE levels in umbilical cord serum. For instance, Okada et al. in 2012^87^ reported lower umbilical cord IgE levels at birth associated with higher maternal PFOA, whereas Wang et al. in 2011^88^ found increased IgE levels in cord blood associated with prenatal PFOA and PFOS. IgE plays an important role in the allergic response; however, baseline total IgE levels may not reliably predict allergic outcomes, particularly because they are not specific to a particular allergen. Additionally, a study by Oulhole et al. 2017 ^95^ found a positive association between PFAS concentrations and basophil counts in 5-year-old infants; while their study did not investigate allergic outcomes, it is well established that basophils play a crucial role in initiating Th2-mediated immunity.^96^ Thus, an enhancement in Th2 responses operates in concert with these innate immune cells, which could lead to the development of adverse allergic outcomes. Type 2 cytokines such as IL-4, IL-5, and IL-13 have been associated with asthma development, and Th2 cells are the primary source of these cytokines.^75,81,82^ Similar to allergic outcomes, evidence for the association between PFAS exposure and asthma has been mixed. Studies reporting negative associations between exposure to PFAS, and the risk of asthma were mainly conducted in children under the age of 10,^91,97^ while those reporting a positive association were conducted in adolescents.^92,94,98^ This emphasizes the importance of considering age and longitudinal analysis when studying immune-related outcomes in children.

Our findings reveal a 0.30 (95% CI: 0.18, 0.41) greater Th2 percentage associated with PFOA at 12 months. While the changes observed in the age-specific percentages of CD4+ T-cells in this study may appear small, these variation sizes have had health implications in previous studies. For instance, an estimated 1.5-2 fold increase in Th2 cells has been noted among infant patients with atopic dermatitis or bronchial asthma compared to healthy controls.^99,100^ Using the mean Th2 percentage of 0.38% (± SD: 0.38) as a reference in our infants (Figure 1F, Supplemental Table 5), an increase of 0.30 indicates an overall 79% increase in Th2 cells. Further, numerous studies have shown that minor changes in the frequency of circulating T follicular helper (cTfh) cells correlate with antigen-specific antibody levels in humans^39,101–103^, at ranges similar to the 18% reduction in Tfh cells we observed. Individuals with higher frequencies of cTfh cells typically exhibit more robust antibody responses; this relationship has been observed in vaccine responses to hepatitis B^103^, yellow fever virus^102^, and influenza^101^, as well as among children infected with malaria^39^.

Our findings provide strong evidence that maternal PFAS exposure affects the infant’s immune system. Differences in CD4+ T cell subpopulations as a function of maternal PFAS exposure support the idea that PFAS are immunotoxic in human infants. As with all research studies using human populations, this study has several strengths, as well as some limitations. One major strength is that this is a longitudinal study with deep immunophenotyping, high-dimensional analysis, and identification of infant CD4+ T-cell subpopulations. Additionally, since only healthy pregnancies and full-term healthy infants were enrolled in the study, we partly removed the potential for congenital-disease-related changes in CD4+ T-cell subpopulations that could influence immunological outcomes. However, this study is not without limitations. The study sample was relatively small (n=200) compared to larger cohort studies.^26,27^ However, even these larger cohort studies that showed negative associations between PFAS and antibody titers did not include in-depth immunophenotyping. These previous studies, therefore, leave a gap in our understanding of the cells responsible for coordinating the antibody response. Moreover, although it is well established that PFAS can cross the placenta^20,104^ and can be detected in the fetus^21^, in the current study, PFAS were only measured during the second trimester of pregnancy and not directly in the infants. Although this is a limitation, this exposure window is a crucial period for potential T-cell programming and development.^2,105^ Human T-cell development begins around the 8^th^ week of pregnancy and continues after birth during infancy; exposures during T-cell development can significantly affect the immune profile of the offspring. ^106^ For example, human cohort studies have linked maternal exposure to pollutants and decreased T-cell-dependent immune defenses in infants and children. ^28,107,108^ Further evidence that developmental exposures shape T-cell function is provided by experimental studies, which provide strong evidence that in-utero exposure can significantly and durably alter T-cell function later in life. ^109^

Participants in the study had lower mean serum concentrations of PFOS, PFOA, and PFNA but higher PFHXS compared to the general population levels reported by NHANES (2017-2018).^61^ The current study provides insights for ongoing risk assessment of PFAS exposures since the legacy PFAS levels are declining over time^61^ and our study suggests immunotoxic impacts in early life, even at these lower levels.

Our results indicate stronger associations between maternal PFAS exposure and CD4+ T-cell subpopulations later during infancy compared to birth. This is likely due to the fact that T-cells are still developing at birth, and infants have not been exposed to the stimuli needed to activate and differentiate T-cell subpopulations. Ongoing PFAS exposure after birth is a key factor to consider. Recently, PFAS has been detected in human breast milk^22,23^ and could represent a continuous source of maternally derived PFAS exposure in the early postnatal life. Rigorous studies to quantify the variation of PFAS in breast milk and formula or other postnatal exposures and immune development are warranted.

In conclusion, we report here an association between in utero exposure to PFAS and decreased percent of Tfh cells and increased percent of Th1, Th2, and Treg cells during the first year of life. These observations support that there are immunomodulatory effects of developmental PFAS exposure. These findings contribute to our understanding of potential mechanisms by which PFAS exposure lowers antibody titers in infants. Moreover, given that imbalances in T-cell subpopulations are associated with other diseases, in-depth profiling of infant T-cells suggests that in utero PFAS exposure may predispose to future immunopathogenesis.

## Data Availability

All data produced in the present study are available upon reasonable request to the authors

## Declaration of competing financial interests (CFI)

The authors declare they have no actual or potential competing financial interests.

## Acknowledgments

Authors would like to acknowledge the support of the mothers and infants participating in UPSIDE-ECHO, UPSIDE-ECHO staff, University of Rochester Flow Cytometry Core, Scheible lab members Adam Geber, Janiret Narvaez Miranda, Dean for their technical assistance. Research reported in this publication was supported by the funding sources: NIH (OD) UH3OD023349, NIH (OD) UG3OD02349, NIH (NIAID) T32AI007285, NIH (NIEHS) P30ES001247, NIH (NIEHS) R01ES036197, NIH (NICHD) R01HD083369, NIH (NCATS) University of Rochester Clinical and Translational Science Award UL1TR002001. The Wynne Center for Family Research. The content is solely the responsibility of the authors and does not necessarily represent the official views of the National Institutes of Health.

