## Supplemental Materials for "In utero exposure to per – and polyfluoroalkyl substances (PFAS) associates with altered human infant T helper cell development"

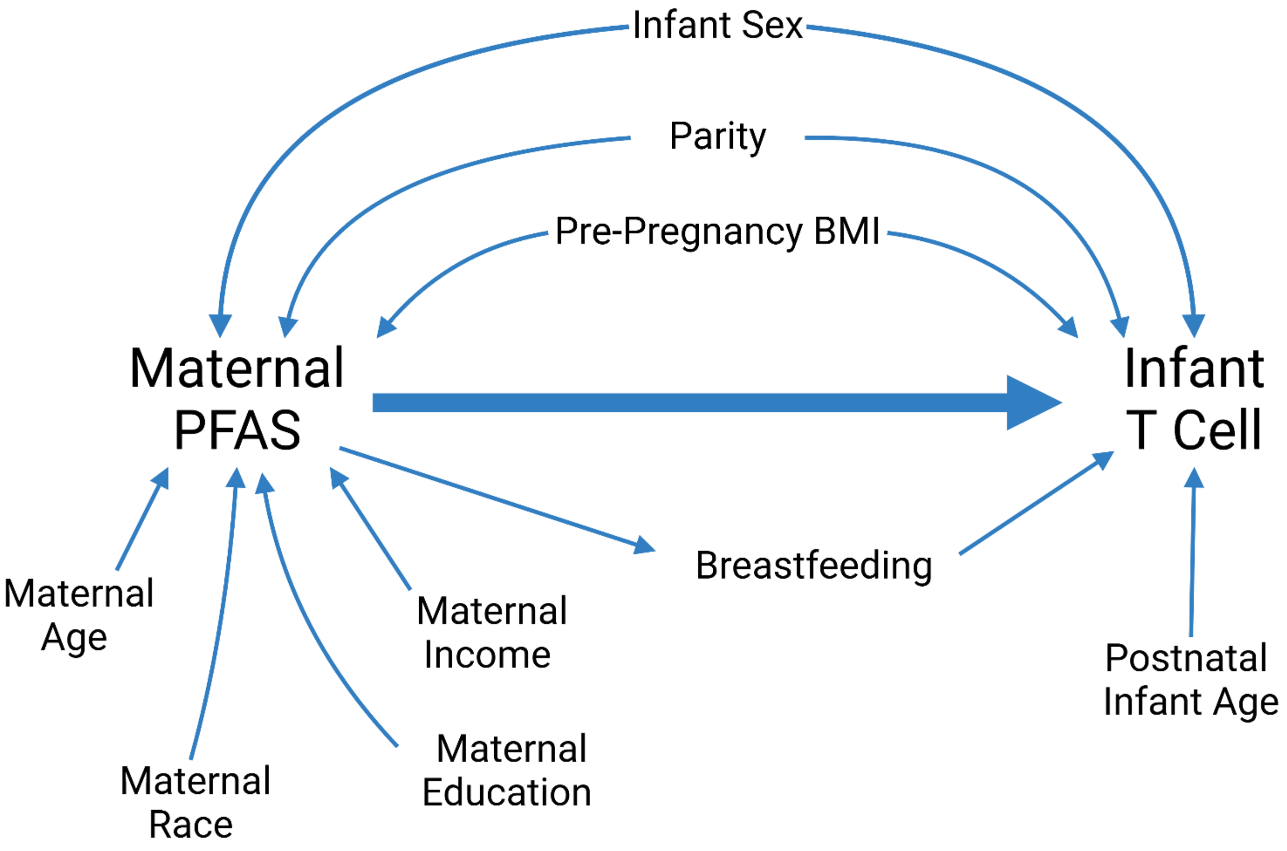


**Supplemental Figure 1:** *Directed acyclic graph (DAG) illustrating the hypothesized relationship between maternal PFAS and infant CD4+ T-cells.* Diagram illustrates the relationship between maternal PFAS (independent), infant CD4+ T-cells (dependent), and potential confounder variables. Arrows indicate hypothesized relationship between each variable.

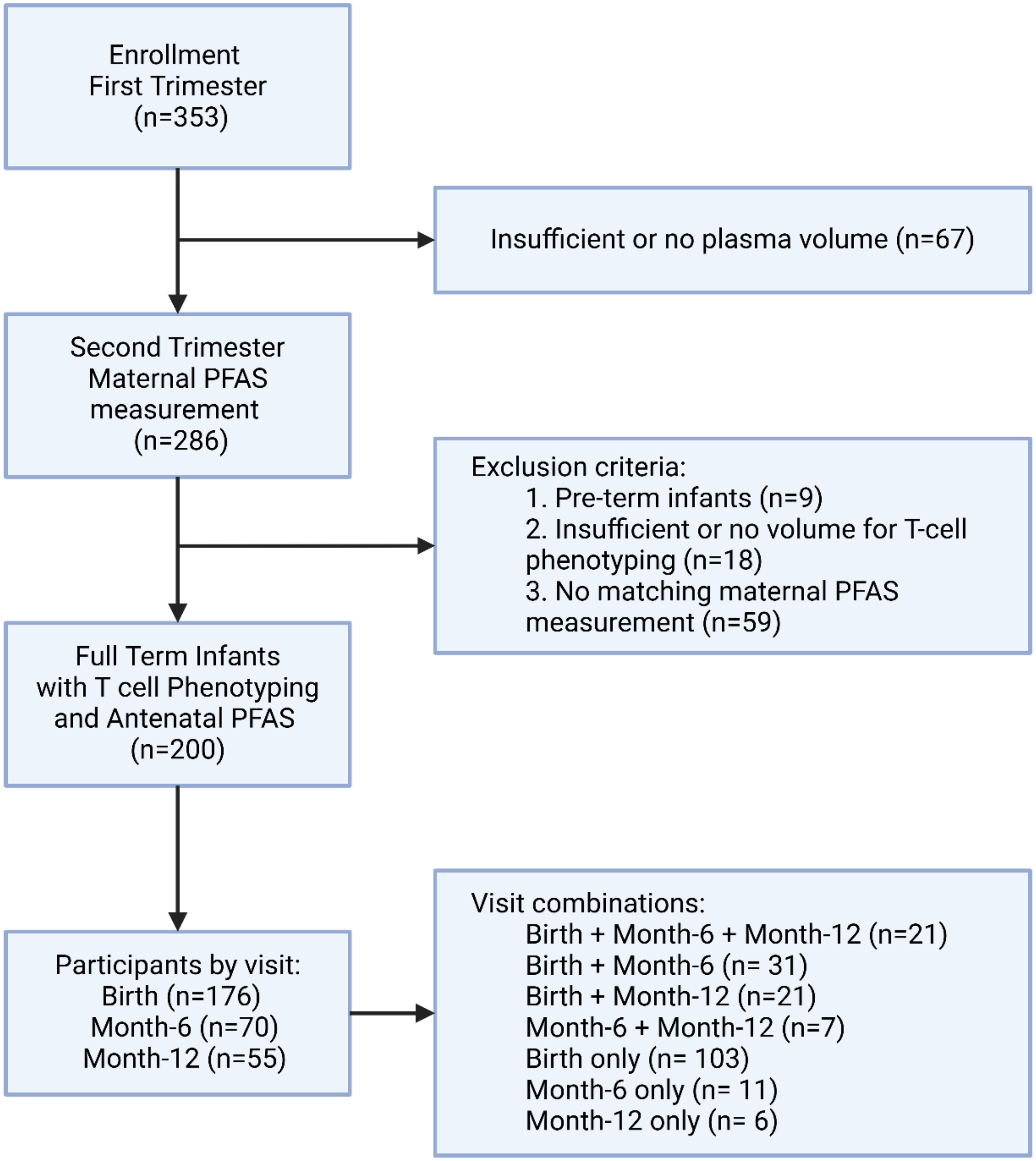


**Supplemental Figure 2:** *UPSIDE-ECHO participant consort diagram for individuals included in the current analysis.*

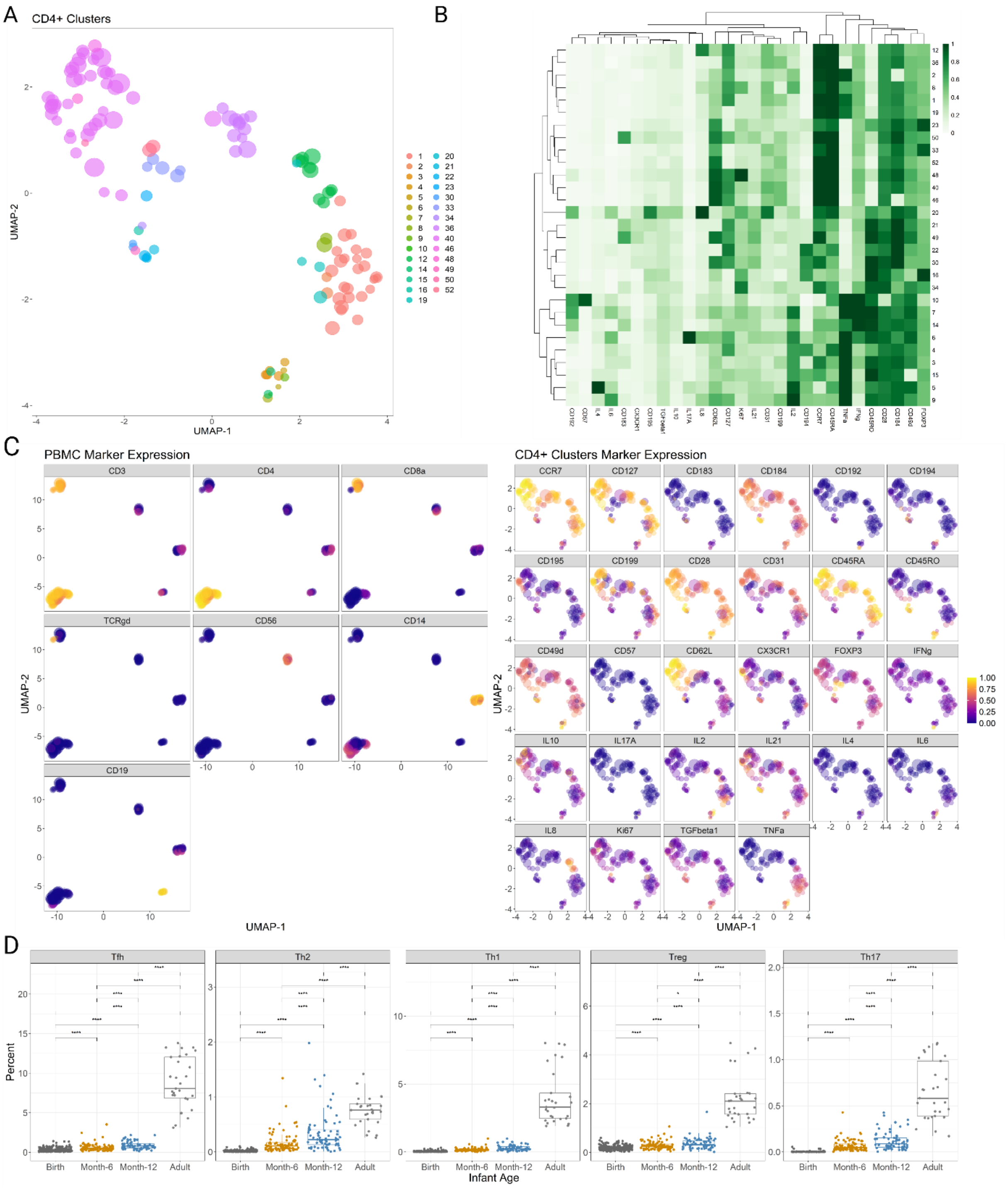


**Supplemental Figure 3:** *Infant T-cell phenotyping.*

(A) Uniform Manifold Approximation and Projection (UMAP) representation of CD4+ FlowSOM nodes colored by sub-cluster number assigned by FlowSOM. (B) Heatmap showing relative marker expression levels on the CD4+ T-cell subpopulations; rows represent individual CD4+ T cell subpopulations identified by FlowSOM, and columns represent each marker. Data were normalized to median marker expression, with color ranging from light green (low expression) to deep green (high expression). Hierarchical clustering, represented by the dendrogram, was applied to rows and columns to determine similarities in CD4+ T-cell subpopulations or marker expression patterns, respectively. (C) UMAP representation of peripheral blood mononuclear cells (PBMC) FlowSOM nodes colored by the expression of PBMC identifying markers: T-cells (CD4+, CD8+, or TCRγδ), Monocytes (CD14+), B cells (CD19+) and NK cells (CD56+) and UMAP representation of CD4+ T cell FlowSOM nodes colored by the expression of each marker included in the panel. Color scale in UMAP represents relative marker expression. (D) Frequency of CD4+ T cells in cord blood at birth and PBMC at 6 months, 12 months, and for adult controls. Each dot on the boxplots denotes data from an individual sample at the indicated age; the y-axis (Percent) is independently scaled for each sub-panel.

**Supplemental Table 1:** Infant immune cell phenotyping panels and parameters.

| Parameter | Target | Antibody clone | Manufacturer | Catalog Number | Metal | Parameter Type | FlowSOM dimensions |
|---|---|---|---|---|---|---|---|
| Time | | | | | | linear (milliseconds) | |
| Event length | | | | | | pulse (duration) | |
| Intercalator 103Rh | Intercalator | | Fluidigm | 201103A | 103Rh | DNA intercalator | PBMC |
| Viability 108Pd* | Viability | | Sigma | in-house§ | 108Pd | viability (DCED) | |
| CD45 106Cd † | CD45 | HI30 | Fluidigm | 3106001B | 106Cd | barcode | 7-choose-3 live-cell barcoding |
| CD45 110Cd † | CD45 | HI30 | Fluidigm | 3110001B | 110Cd | barcode | 7-choose-3 live-cell barcoding |
| CD45 111Cd † | CD45 | HI30 | Fluidigm | 3111001B | 111Cd | barcode | 7-choose-3 live-cell barcoding |
| CD45 112Cd † | CD45 | HI30 | Fluidigm | 3112001B | 112Cd | barcode | 7-choose-3 live-cell barcoding |
| CD45 113Cd † | CD45 | HI30 | Fluidigm | 3113001B | 113Cd | barcode | 7-choose-3 live-cell barcoding |
| CD45 113In* | CD45 | HI30 | BioLegend | 304045** | 113In | barcode | 6-choose-3 live-cell barcoding |
| CD45 114Cd † | CD45 | HI30 | Fluidigm | 3114001B | 114Cd | barcode | 7-choose-3 live-cell barcoding |
| CD45 115In* | CD45 | HI30 | BioLegend | 304045** | 115In | barcode | 6-choose-3 live-cell barcoding |
| CD45 116Cd † | CD45 | HI30 | Fluidigm | 3116001B | 116Cd | barcode | 7-choose-3 live-cell barcoding |
| Beads 140Ce | Beads | | Fluidigm | 201078 | 140Ce | normalization bead (EQ) | |
| CD49d 141Pr | CD49d | 9F10 | Fluidigm | 3141004B | 141Pr | surface | T-cells |
| CD19 142Nd | CD19 | HIB19 | Fluidigm | 3142001B | 142Nd | surface | PBMC |
| IL2 143Nd | IL2 | MQ1-17H12 | BioLegend | 500339** | 143Nd | intracellular | T-cells |
| CD31 144Nd | CD31 | WM59 | Fluidigm | 3144023B | 144Nd | surface | T-cells |
| CD4 145Nd | CD4 | RPA-T4 | Fluidigm | 3145001B | 145Nd | surface | PBMC |
| CD14 146Nd | CD14 | M5E2 | BioLegend | 301843** | 146Nd | surface | PBMC |
| IL4 147Sm | IL4 | MP4-25D2 | BioLegend | 500829** | 147Sm | intracellular | T-cells |
| IL17A 148Nd | IL17A | BL168 | Fluidigm | 3148008B | 148Nd | intracellular | T-cells |
| CD56 149Sm | CD56 | NCAM16.2 | Fluidigm | 3149021B | 149Sm | surface | PBMC |
| IL8 150Nd | IL8 | E8N1 | BioLegend | 511402** | 150Nd | intracellular | T-cells |
| CD69 151Eu † | CD69 | FN50 | BioLegend | 310939** | 151Eu | intracellular | not used for clustering |
| TNFa 152Sm | TNFa | Mab11 | Fluidigm | 3152002B | 152Sm | intracellular | T-cells |
| CD62L 153Eu | CD62L | DREG-56 | Fluidigm | 3153004B | 153Eu | surface | T-cells |
| IL6 154Sm | IL6 | MQ2-13A5 | Fluidigm | 3154011B | 154Sm | intracellular | T-cells |
| TCRgd 155Gd † | TCRgd | B1 | BioLegend | 331202** | 155Gd | surface | PBMC |
| CD183 156Gd | CD183 | G025H7 | Fluidigm | 3156004B | 156Gd | surface | T-cells |
| TGFbeta1 158Gd | TGFbeta1 | TW4-6H10 | BioLegend | discontinued** | 158Gd | intracellular | T-cells |

| | | | | | | | |
|---|---|---|---|---|---|---|---|
| FOXP3 159Tb | FoxP3 | 259D/C7 | Fluidigm | 3159028A | 159Tb | intracellular | T-cells |
| CD28 160Gd | CD28 | CD28.2 | Fluidigm | 3160003B | 160Gd | surface | T-cells |
| Ki67 161Dy | Ki67 | B56 | Fluidigm | 3161007B | 161Dy | intracellular | T-cells |
| CD8a 162Dy | CD8a | RPA-T8 | Fluidigm | 3162015B | 162Dy | surface | PBMC |
| CD192 163Dy | CD192 | K036C2 | BioLegend | 357202** | 163Dy | surface | T-cells |
| IFNg 164Dy | IFNg | B27 | BioLegend | 506521** | 164Dy | intracellular | T-cells |
| CD45RO 165Ho | CD45RO | UCHL1 | Fluidigm | 3165011B | 165Ho | surface | T-cells |
| IL10 166Er | IL10 | JES3-9D7 | Fluidigm | 3166008B | 166Er | intracellular | T-cells |
| CCR7 167Er | CCR7 | G043H7 | Fluidigm | 3167009A | 167Er | surface | T-cells |
| CD199 168Er | CD199 | L053E8 | Fluidigm | 3168011A | 168Er | surface | T-cells |
| CD45RA 169Tm | CD45RA | HI100 | Fluidigm | 3169008B | 169Tm | surface | T-cells |
| CD3 170Er | CD3 | UCHT1 | Fluidigm | 3170001B | 170Er | intracellular | PBMC |
| CD195 171Yb | CD195 | NP-6G4 | Fluidigm | 3171017A | 171Yb | surface | T-cells |
| CX3CR1 172Yb | CX3CR1 | 2A9-1 | Fluidigm | 3172017B | 172Yb | surface | T-cells |
| CD184 173Yb | CD184 | 12G5 | Fluidigm | 3173001B | 173Yb | surface | T-cells |
| IL21 174Yb | IL21 | 3A3-N2 | BioLegend | 513009** | 174Yb | intracellular | T-cells |
| CD194 175Lu | CD194 | L291H4 | Fluidigm | 3175035A | 175Lu | surface | T-cells |
| CD127 176Yb | CD127 | A019D5 | Fluidigm | 3176004B | 176Yb | surface | T-cells |
| CD45 194Pt* | CD45 | HI30 | BioLegend | 304045¶ | 194Pt | barcode | 6-choose-3 live-cell barcoding |
| Viability 194Pt † | Viability | | Fluidigm | 201194 | 194Pt | viability (Cisplatin) | |
| CD45 195Pt* | CD45 | HI30 | BioLegend | 304045¶ | 195Pt | barcode | 6-choose-3 live-cell barcoding |
| CD45 196Pt* | CD45 | HI30 | BioLegend | 304045¶ | 196Pt | barcode | 6-choose-3 live-cell barcoding |
| CD45 198Pt* | CD45 | HI30 | BioLegend | 304045¶ | 198Pt | barcode | 6-choose-3 live-cell barcoding |
| CD57 209Bi | CD57 | HCD57 | Fluidigm | special order | 209Bi | surface | T-cells |
| Center | | | | | | pulse (Gaussian) | |
| Offset | | | | | | pulse (Gaussian) | |
| Width | | | | | | pulse (Gaussian) | |
| Residual | | | | | | pulse (Gaussian) | |

* Initial panel parameters; † Revised panel parameters; **Purified antibodies were conjugated to metal isotopes using a Maxpar X8 labeling kit (Fluidigm/Standard BioTools); § Palladium-based viability stain [50]; ¶ Platinum-conjugated antibodies [52]; PBMC: peripheral blood mononuclear cells

**Supplemental Table 2:** Infant and maternal characteristics of the UPSIDE-ECHO population by infant age**.**

| **Characteristic** | **Total** | **Birth** | **Month-6** | **Month-12** |
|---|---|---|---|---|
| N (%) | 200(100) | 176(88) | 70(35) | 55(27.5) |
| | **Mean (SD)** | **Mean (SD)** | **Mean (SD)** | **Mean (SD)** |
| Gestational Age (weeks) | 39.74±1.11 | 39.71±1.10 | 39.85±1.07 | 39.71±1.13 |
| Birth Weight (grams) | 3411.39±464.49 | 3403.82±451.50 | 3519.27±450.73 | 3423.42±472.00 |
| | **N (%)** | **N (%)** | **N (%)** | **N (%)** |
| Maternal Age | | | | |
| 18-25 | 41(20.5) | 39(22.16) | 9(12.86) | 6(10.91) |
| 26-35 | 142(71) | 121(68.75) | 57(81.43) | 46(83.64) |
| 36+ | 17(8.5) | 16(9.09) | 4(5.71) | 3(5.45) |
| Pre-pregnancy BMI | | | | |
| Underweight | 1(0.5) | 0(0) | 1(1.43) | 0(0) |
| Normal Weight | 90(45) | 78(44.32) | 33(47.14) | 23(41.82) |
| Overweight | 50(25) | 46(26.14) | 18(25.71) | 14(25.45) |
| Obese | 59(29.5) | 52(29.55) | 18(25.71) | 18(32.73) |
| Annual Income | | | | |
| Lower Class | 41(20.5) | 36(20.45) | 12(17.14) | 6(10.91) |
| Lower-Middle Class | 48(24) | 43(24.43) | 20(28.57) | 11(20) |
| Middle Class | 40(20) | 36(20.45) | 12(17.14) | 15(27.27) |
| Upper-Middle Class | 34(17) | 29(16.48) | 14(20) | 12(21.82) |
| Upper Class | 19(9.5) | 14(7.95) | 9(12.86) | 7(12.73) |
| Not Reported | 18(9) | 18(10.23) | 3(4.29) | 4(7.27) |
| Parity | | | | |
| 0 | 69(34.5) | 59(33.52) | 21(30) | 17(30.91) |
| 1 | 78(39) | 72(40.91) | 28(40) | 21(38.18) |
| 2+ | 53(26.5) | 45(25.57) | 21(30) | 17(30.91) |
| Infant Sex | | | | |
| Male | 105(52.5) | 92(52.27) | 44(62.86) | 34(61.82) |
| Female | 95(47.5) | 84(47.73) | 26(37.14) | 21(38.18) |
| Mode of Delivery | | | | |
| Vaginal | 153(76.5) | 138(78.41) | 53(75.71) | 46(83.64) |
| Caesarian | 47(23.5) | 38(21.59) | 17(24.29) | 9(16.36) |
| Maternal Race | | | | |

| | | | | |
|---|---|---|---|---|
| White | 125(62.5) | 106(60.23) | 50(71.43) | 39(70.91) |
| Asian | 40(20) | 36(20.45) | 12(17.14) | 12(21.82) |
| Black | 16(8) | 16(9.09) | 3(4.29) | 1(1.82) |
| Hispanic | 9(4.5) | 9(5.11) | 2(2.86) | 2(3.64) |
| Other | 10(5) | 9(5.11) | 3(4.29) | 1(1.82) |
| Maternal Education | | | | |
| HS or Less | 64(32) | 57(32.39) | 22(31.43) | 11(20) |
| Some college | 31(15.5) | 27(15.34) | 5(7.14) | 8(14.55) |
| Bachelors | 54(27) | 52(29.55) | 20(28.57) | 16(29.09) |
| Postgraduate | 50(25) | 39(22.16) | 23(32.86) | 20(36.36) |
| Not Reported | 1(0.5) | 1(0.57) | 0(0) | 0(0) |
| Medicaid During Pregnancy | | | | |
| No | 114(57) | 99(56.25) | 44(62.86) | 37(67.27) |
| Yes | 86(43) | 77(43.75) | 26(37.14) | 18(32.73) |

BMI: Body Mass Index, SD: Standard deviation, HS: High School.

**Supplemental Table 3**: Distribution of the 14 perfluoroalkyl substances (PFAS)(ng/mL) measured in maternal plasma in the UPSIDE-ECHO population.

| PFAS | N | Samples >LOD | Percent detected | LOD | Geometric Mean | Geometric SD | Min | 10th | 25th | Median | 75th | 90th | Max |
|---|---|---|---|---|---|---|---|---|---|---|---|---|---|
| PFOS | 200 | 200 | 100 | 0.020 | 2.478 | 1.653 | 0.611 | 1.331 | 1.828 | 2.510 | 3.215 | 4.188 | 21.304 |
| PFOA | 200 | 200 | 100 | 0.020 | 0.572 | 1.799 | 0.059 | 0.297 | 0.400 | 0.585 | 0.827 | 1.078 | 5.448 |
| PFNA | 200 | 200 | 100 | 0.032 | 0.26 | 1.821 | 0.063 | 0.144 | 0.170 | 0.239 | 0.353 | 0.489 | 2.874 |
| PFHXS | 200 | 200 | 100 | 0.020 | 1.782 | 1.549 | 0.560 | 1.042 | 1.380 | 1.754 | 2.422 | 3.166 | 7.166 |
| PFDA | 200 | 160 | 80 | 0.020 | 0.07 | 2.014 | <LOD | <LOD | 0.028 | 0.053 | 0.092 | 0.142 | 1.042 |
| PFPEA | 200 | 120 | 60 | 0.0224 | NC | NC | <LOD | <LOD | <LOD | 0.042 | 0.159 | 0.297 | 1.088 |
| PFUNDA | 200 | 120 | 60 | 0.020 | NC | NC | <LOD | <LOD | <LOD | 0.031 | 0.074 | 0.112 | 0.366 |
| NMFOSAA | 200 | 58 | 29 | 0.032 | NC | NC | <LOD | <LOD | <LOD | <LOD | 0.039 | 0.084 | 0.543 |
| PFBS | 200 | 19 | 9.5 | 0.020 | NC | NC | <LOD | <LOD | <LOD | <LOD | <LOD | <LOD | 0.569 |
| PFDODA | 200 | 15 | 7.5 | 0.0224 | NC | NC | <LOD | <LOD | <LOD | <LOD | <LOD | <LOD | 0.11 |
| PFHPA | 200 | 13 | 6.5 | 0.0224 | NC | NC | <LOD | <LOD | <LOD | <LOD | <LOD | <LOD | 0.162 |
| PFHXA | 200 | 10 | 5 | 0.020 | NC | NC | <LOD | <LOD | <LOD | <LOD | <LOD | <LOD | 0.188 |
| NETFOSAA | 200 | 7 | 3.5 | 0.032 | NC | NC | <LOD | <LOD | <LOD | <LOD | <LOD | <LOD | 0.074 |
| PFOSA | 200 | 0 | 0 | 0.02 | NC | NC | <LOD | <LOD | <LOD | <LOD | <LOD | <LOD | 0 |

PFOS: perfluorooctanesulfonic acid, PFOA: perfluorooctanoic acid, PFHxS: perfluorohexane sulfonic acid, PFNA: perfluorononanoic acid, PFDA: perfluorodecanoic acid, PFPEA: perfluoropentanoic acid, PFUNDA: perfluoroundecanoic acid, NMFOSAA: N-Methyl perfluorooctane sulfonamidoacetic acid, PFBS: perfluorobutane sulfonic acid, PFDODA: perfluorodecanoic acid, PFHPA: perfluoroheptanoic acid, PFHXA: perfluorohexanoic acid, NETFOSAA: N-Ethyl perfluorooctane sulfonamidoacetic acid, PFOSA: perfluorooctane sulfonamide,SD: Standard Deviation, Min: minimum, Max: maximum, LOD: limit of detection, NC: Not computed.

**Supplemental Table 4:** Determinants of maternal exposures to perfluoroalkyl substances (PFAS)(ng/mL) in the UPSIDE-ECHO population.

| ***Categorical Covariates*** | | | | | | | | | | |
|---|---|---|---|---|---|---|---|---|---|---|
| ***Parity*** | | | | | | | | | | |
| **PFAS** | **Parity** | **Mean±SD** | **Group1** | **Group2** | **N1** | **N2** | **Estimate (95%CI)** | **Statistic** | **P-value** | **Adjusted P-value** |
| PFOS | 0 | 3.174±1.86 | 0 | 1 | 69 | 78 | 0.367( 0.00,0.73) | 3199 | 0.049 | 0.098 |
| | 1 | 2.737±1.81 | 0 | 2+ | 69 | 53 | 0.611( 0.21,0.97) | 2401 | 0.003 | 0.009 |
| | 2+ | 2.661±2.78 | 1 | 2+ | 78 | 53 | 0.233(-0.10,0.57) | 2365 | 0.163 | 0.163 |
| PFOA | 0 | 0.923±0.69 | 0 | 1 | 69 | 78 | 0.210( 0.12,0.31) | 3730 | 0.000 | 0.000 |
| | 1 | 0.613±0.30 | 0 | 2+ | 69 | 53 | 0.342( 0.26,0.44) | 3099 | 0.000 | 0.000 |
| | 2+ | 0.469±0.30 | 1 | 2+ | 78 | 53 | 0.138( 0.07,0.22) | 2797 | 0.001 | 0.001 |
| PFNA | 0 | 0.366±0.32 | 0 | 1 | 69 | 78 | 0.042( 0.00,0.08) | 3186 | 0.055 | 0.055 |
| | 1 | 0.308±0.31 | 0 | 2+ | 69 | 53 | 0.082( 0.04,0.13) | 2568 | 0.000 | 0.000 |
| | 2+ | 0.316±0.47 | 1 | 2+ | 78 | 53 | 0.045( 0.01,0.08) | 2598 | 0.013 | 0.026 |
| PFHXS | 0 | 2.005±1.02 | 0 | 1 | 69 | 78 | -0.079(-0.34,0.17) | 2540 | 0.558 | 0.558 |
| | 1 | 2.096±0.96 | 0 | 2+ | 69 | 53 | 0.233(-0.06,0.47) | 2127 | 0.124 | 0.248 |
| | 2+ | 1.706±0.64 | 1 | 2+ | 78 | 53 | 0.313( 0.05,0.55) | 2564 | 0.020 | 0.060 |
| ***Infant Sex*** | | | | | | | | | | |
| **PFAS** | **Infant Sex** | **Mean±SD** | **Group1** | **Group2** | **N1** | **N2** | **Estimate (95%CI)** | **Statistic** | **P-value** | |
| PFOS | Male | 3.015±1.98 | Male | Female | 105 | 95 | 0.305( 0.01,0.60) | 5807 | 0.045 | |
| | Female | 2.705±2.28 | | | | | | | | |
| PFOA | Male | 0.740±0.60 | Male | Female | 105 | 95 | 0.096( 0.00,0.18) | 5825 | 0.041 | |
| | Female | 0.617±0.37 | | | | | | | | |
| PFNA | Male | 0.310±0.21 | Male | Female | 105 | 95 | 0.022(-0.01,0.06) | 5557 | 0.164 | |
| | Female | 0.352±0.48 | | | | | | | | |
| PFHXS | Male | 2.015±0.93 | Male | Female | 105 | 95 | 0.118(-0.09,0.33) | 5435 | 0.275 | |
| | Female | 1.901±0.91 | | | | | | | | |
| ***Mode of Delivery*** | | | | | | | | | | |
| **PFAS** | **Mode of Delivery** | **Mean±SD** | **Group1** | **Group2** | **N1** | **N2** | **Estimate (95%CI)** | **Statistic** | **P-value** | |
| PFOS | CS | 2.985±1.77 | CS | Vaginal | 47 | 153 | 0.233(-0.17,0.62) | 4002 | 0.242 | |
| | Vaginal | 2.832±2.23 | | | | | | | | |
| PFOA | CS | 0.654±0.37 | CS | Vaginal | 47 | 153 | -0.003(-0.10,0.10) | 3576 | 0.956 | |
| | Vaginal | 0.690±0.54 | | | | | | | | |
| PFNA | CS | 0.276±0.16 | CS | Vaginal | 47 | 153 | -0.017(-0.06,0.02) | 3277 | 0.360 | |
| | Vaginal | 0.347±0.40 | | | | | | | | |

| | | | | | | | | | | |
|---|---|---|---|---|---|---|---|---|---|---|
| PFHXS | CS<br>Vaginal | 2.002±1.13<br>1.949±0.85 | CS | Vaginal | 47 | 153 | -0.024(-0.28,0.24) | 3535 | 0.862 | |
| ***Maternal Race*** | | | | | | | | | | |
| **PFAS** | **Race** | **Mean±SD** | **Group1** | **Group2** | **N1** | **N2** | **Estimate (95%CI)** | **Statistic** | **P-value** | **Adjusted P-value** |
| PFOS | White | 3.126±2.54 | White | Black | 125 | 16 | -0.021(-0.63, 0.60) | 985 | 0.925 | 1.000 |
| | Black | 2.700±1.04 | White | Hispanic | 125 | 9 | 0.724( 0.05, 1.42) | 800 | 0.035 | 0.316 |
| | Hispanic | 2.010±0.96 | White | Asian | 125 | 40 | 0.406( 0.07, 0.75) | 3114 | 0.020 | 0.197 |
| | Asian | 2.323±0.98 | White | Other | 125 | 10 | -0.116(-0.95, 0.66) | 593 | 0.791 | 1.000 |
| | Other | 2.863±1.10 | Black | Hispanic | 16 | 9 | 0.789(-0.18, 1.55) | 98 | 0.152 | 0.876 |
| | | | Black | Asian | 16 | 40 | 0.433(-0.19, 1.06) | 401 | 0.146 | 0.876 |
| | | | Black | Other | 16 | 10 | -0.156(-1.16, 0.85) | 71 | 0.660 | 1.000 |
| | | | Hispanic | Asian | 9 | 40 | -0.387(-0.97, 0.31) | 132 | 0.224 | 0.896 |
| | | | Hispanic | Other | 9 | 10 | -0.893(-2.08, 0.02) | 22 | 0.065 | 0.522 |
| | | | Asian | Other | 40 | 10 | -0.515(-1.39, 0.20) | 136 | 0.125 | 0.875 |
| PFOA | White | 0.714±0.39 | White | Black | 125 | 16 | -0.010(-0.16, 0.16) | 978 | 0.889 | 1.000 |
| | Black | 0.678±0.26 | White | Hispanic | 125 | 9 | 0.242( 0.06, 0.45) | 849 | 0.011 | 0.088 |
| | Hispanic | 0.415±0.20 | White | Asian | 125 | 40 | 0.173( 0.08, 0.28) | 3445 | 0.000 | 0.003 |
| | Asian | 0.591±0.81 | White | Other | 125 | 10 | -0.109(-0.32, 0.14) | 529 | 0.422 | 1.000 |
| | Other | 0.893±0.64 | Black | Hispanic | 16 | 9 | 0.253( 0.05, 0.49) | 116 | 0.012 | 0.088 |
| | | | Black | Asian | 16 | 40 | 0.203( 0.05, 0.35) | 457 | 0.012 | 0.088 |
| | | | Black | Other | 16 | 10 | -0.091(-0.36, 0.19) | 66 | 0.484 | 1.000 |
| | | | Hispanic | Asian | 9 | 40 | -0.076(-0.22, 0.10) | 151 | 0.468 | 1.000 |
| | | | Hispanic | Other | 9 | 10 | -0.329(-0.65,-0.10) | 12 | 0.006 | 0.051 |
| | | | Asian | Other | 40 | 10 | -0.275(-0.46,-0.05) | 100 | 0.014 | 0.088 |
| PFNA | White | 0.317±0.31 | White | Black | 125 | 16 | -0.013(-0.07, 0.05) | 934 | 0.670 | 1.000 |
| | Black | 0.276±0.09 | White | Hispanic | 125 | 9 | 0.079( 0.02, 0.15) | 848 | 0.011 | 0.090 |
| | Hispanic | 0.176±0.06 | White | Asian | 125 | 40 | 0.004(-0.04, 0.05) | 2551 | 0.848 | 1.000 |
| | Asian | 0.424±0.57 | White | Other | 125 | 10 | -0.077(-0.16, 0.01) | 433 | 0.108 | 0.648 |
| | Other | 0.337±0.11 | Black | Hispanic | 16 | 9 | 0.096( 0.04, 0.17) | 123 | 0.003 | 0.025 |
| | | | Black | Asian | 16 | 40 | 0.031(-0.05, 0.08) | 358 | 0.500 | 1.000 |
| | | | Black | Other | 16 | 10 | -0.069(-0.16, 0.03) | 56 | 0.220 | 0.880 |
| | | | Hispanic | Asian | 9 | 40 | -0.061(-0.16, 0.00) | 97 | 0.032 | 0.220 |
| | | | Hispanic | Other | 9 | 10 | -0.183(-0.26,-0.04) | 7 | 0.001 | 0.010 |
| | | | Asian | Other | 40 | 10 | -0.078(-0.18, 0.04) | 137 | 0.131 | 0.655 |
| PFHXS | White | 2.078±0.99 | White | Black | 125 | 16 | 0.211(-0.16, 0.60) | 1173 | 0.262 | 1.000 |

| | | | | | | | | | | |
|---|---|---|---|---|---|---|---|---|---|---|
| | Black | 1.776±0.64 | White | Hispanic | 125 | 9 | 0.340(-0.17, 0.94) | 720 | 0.163 | 1.000 |
| | Hispanic | 1.610±0.66 | White | Asian | 125 | 40 | 0.215(-0.08, 0.50) | 2879.5 | 0.150 | 1.000 |
| | Asian | 1.867±0.87 | White | Other | 125 | 10 | 0.413( 0.06, 0.89) | 887 | 0.028 | 0.280 |
| | Other | 1.489±0.29 | Black | Hispanic | 16 | 9 | 0.066(-0.40, 0.78) | 76 | 0.846 | 1.000 |
| | | | Black | Asian | 16 | 40 | -0.018(-0.43, 0.39) | 317 | 0.964 | 1.000 |
| | | | Black | Other | 16 | 10 | 0.182(-0.20, 0.72) | 93 | 0.517 | 1.000 |
| | | | Hispanic | Asian | 9 | 40 | -0.145(-0.72, 0.41) | 154 | 0.517 | 1.000 |
| | | | Hispanic | Other | 9 | 10 | 0.116(-0.40, 0.56) | 48 | 0.842 | 1.000 |
| | | | Asian | Other | 40 | 10 | 0.178(-0.20, 0.64) | 243 | 0.308 | 1.000 |
| ***Maternal Education*** | | | | | | | | | | |
| **PFAS** | **Education** | **Mean±SD** | **Group1** | **Group2** | **N1** | **N2** | **Estimate (95%CI)** | **Statistic** | **P-value** | **Adjusted P-value** |
| PFOS | HS or Less | 2.429±1.24 | HS or Less | Postgraduate | 64 | 50 | -0.691(-1.09,-0.29) | 1011 | 0.001 | 0.005 |
| | Some college | 2.456±1.14 | Some college | Bachelors | 31 | 54 | -0.166(-0.70, 0.33) | 758 | 0.474 | 0.948 |
| | Bachelors | 2.888±2.11 | HS or Less | Bachelors | 64 | 54 | -0.266(-0.60, 0.08) | 1454 | 0.140 | 0.420 |
| | Postgraduate | 3.645±3.14 | Bachelors | Postgraduate | 54 | 50 | -0.426(-0.86, 0.00) | 1049 | 0.051 | 0.202 |
| | | | HS or Less | Some college | 64 | 31 | -0.078(-0.56, 0.36) | 950 | 0.742 | 0.948 |
| | | | Some college | Postgraduate | 31 | 50 | -0.617(-1.20,-0.10) | 541 | 0.023 | 0.116 |
| PFOA | HS or Less | 0.577±0.29 | HS or Less | Postgraduate | 64 | 50 | -0.210(-0.33,-0.09) | 996 | 0.001 | 0.003 |
| | Some college | 0.605±0.26 | Some college | Bachelors | 31 | 54 | 0.023(-0.11, 0.14) | 877 | 0.718 | 1.000 |
| | Bachelors | 0.733±0.79 | HS or Less | Bachelors | 64 | 54 | -0.015(-0.12, 0.09) | 1677 | 0.785 | 1.000 |
| | Postgraduate | 0.815±0.43 | Bachelors | Postgraduate | 54 | 50 | -0.188(-0.31,-0.04) | 970 | 0.014 | 0.068 |
| | | | HS or Less | Some college | 64 | 31 | -0.042(-0.16, 0.08) | 918 | 0.560 | 1.000 |
| | | | Some college | Postgraduate | 31 | 50 | -0.162(-0.33,-0.01) | 556 | 0.034 | 0.135 |
| PFNA | HS or Less | 0.333±0.44 | HS or Less | Postgraduate | 64 | 50 | -0.064(-0.12,-0.02) | 1133 | 0.008 | 0.046 |
| | Some college | 0.255±0.10 | Some college | Bachelors | 31 | 54 | -0.013(-0.07, 0.03) | 773 | 0.562 | 0.946 |
| | Bachelors | 0.308±0.23 | HS or Less | Bachelors | 64 | 54 | -0.022(-0.07, 0.02) | 1517 | 0.256 | 0.768 |
| | Postgraduate | 0.396±0.45 | Bachelors | Postgraduate | 54 | 50 | -0.040(-0.09, 0.01) | 1130 | 0.153 | 0.612 |
| | | | HS or Less | Some college | 64 | 31 | -0.013(-0.05, 0.03) | 901 | 0.473 | 0.946 |
| | | | Some college | Postgraduate | 31 | 50 | -0.055(-0.11, 0.00) | 568 | 0.045 | 0.224 |
| PFHXS | HS or Less | 1.712±0.77 | HS or Less | Postgraduate | 64 | 50 | -0.493(-0.77,-0.20) | 1026 | 0.001 | 0.006 |
| | Some college | 1.917±0.85 | Some college | Bachelors | 31 | 54 | -0.069(-0.38, 0.31) | 799 | 0.732 | 0.732 |
| | Bachelors | 1.930±0.74 | HS or Less | Bachelors | 64 | 54 | -0.216(-0.48, 0.03) | 1420 | 0.097 | 0.438 |
| | Postgraduate | 2.325±1.19 | Bachelors | Postgraduate | 54 | 50 | -0.232(-0.58, 0.05) | 1087 | 0.088 | 0.438 |
| | | | HS or Less | Some college | 64 | 31 | -0.191(-0.50, 0.13) | 845 | 0.245 | 0.490 |
| | | | Some college | Postgraduate | 31 | 50 | -0.313(-0.68, 0.08) | 616 | 0.124 | 0.438 |

| *Medicaid During Pregnancy* | | | | | | | | | |
|---|---|---|---|---|---|---|---|---|---|
| **PFAS** | **Medicaid** | **Mean±SD** | **Group1** | **Group2** | **N1** | **N2** | **Estimate (95%CI)** | **Statistic** | **P-value** |
| PFOS | No<br>Yes | 3.285±2.60<br>2.315±1.05 | No | Yes | 114 | 86 | 0.604(0.31,0.89) | 6529 | 0.000 |
| PFOA | No<br>Yes | 0.772±0.61<br>0.563±0.27 | No | Yes | 114 | 86 | 0.133(0.05,0.23) | 6104 | 0.003 |
| PFNA | No<br>Yes | 0.337±0.34<br>0.321±0.39 | No | Yes | 114 | 86 | 0.030(0.00,0.07) | 5653 | 0.064 |
| PFHXS | No<br>Yes | 2.122±0.98<br>1.748±0.79 | No | Yes | 114 | 86 | 0.329(0.12,0.54) | 6145 | 0.002 |

| **Continuous Variables** | | | |
|---|---|---|---|
| **PFAS** | **Covariate** | **Spearman's Rho** | **P-value** |
| | ***Maternal Age*** | | |
| PFOS | | 0.118 | 0.096 |
| PFOA | | 0.110 | 0.120 |
| PFNA | | 0.006 | 0.937 |
| PFHXS | | 0.026 | 0.716 |
| | ***Pre-pregnancy BMI*** | | |
| PFOS | | -0.211 | 0.003 |
| PFOA | | -0.014 | 0.844 |
| PFNA | | -0.070 | 0.328 |
| PFHXS | | -0.227 | 0.001 |
| | ***Annual Income*** | | |
| PFOS | | 0.244 | 0.001 |
| PFOA | | 0.271 | 0.000 |
| PFNA | | 0.132 | 0.075 |
| PFHXS | | 0.198 | 0.007 |
| | ***Birth Weight*** | | |
| PFOS | | 0.034 | 0.638 |
| PFOA | | 0.041 | 0.568 |
| PFNA | | -0.076 | 0.286 |
| PFHXS | | 0.109 | 0.123 |

PFOS: perfluorooctanesulfonic acid, PFOA: perfluorooctanoic acid, PFHxS: perfluorohexane sulfonic acid, PFNA: perfluorononanoic acid, PFDA: perfluorodecanoic acid, SD: Standard Deviation, CI: confidence interval, HS: high school, CS: cesarean section, P- values adjusted for multiple comparisons using the Holm-Bonferroni method.

**Supplemental Table 5**: Distribution of CD4+ T-cells by age.

| Cell Type | Visit | N (%) | Mean ±SD | Min | Q1 | Q3 | Max |
|---|---|---|---|---|---|---|---|
| Tfh | Birth | 176(88) | 0.34 ± 0.23 | 0.06 | 0.17 | 0.47 | 1.41 |
| | Month-6 | 70(35) | 0.61 ± 0.36 | 0.20 | 0.35 | 0.77 | 2.45 |
| | Month-12 | 55(27.5) | 0.96 ± 0.46 | 0.38 | 0.64 | 1.11 | 2.17 |
| | Adult | 7(100) | 8.35 ± 3.41 | 3.60 | 6.24 | 10.68 | 12.87 |
| Th2 | Birth | 176(88) | 0.02 ± 0.02 | 0.00 | 0.01 | 0.02 | 0.10 |
| | Month-6 | 70(35) | 0.17 ± 0.21 | 0.01 | 0.06 | 0.20 | 1.34 |
| | Month-12 | 55(27.5) | 0.38 ± 0.38 | 0.02 | 0.15 | 0.47 | 1.98 |
| | Adult | 7(100) | 0.75 ± 0.32 | 0.36 | 0.55 | 0.89 | 1.30 |
| Th1 | Birth | 176(88) | 0.04 ± 0.04 | 0.00 | 0.02 | 0.05 | 0.31 |
| | Month-6 | 70(35) | 0.18 ± 0.14 | 0.02 | 0.11 | 0.21 | 0.98 |
| | Month-12 | 55(27.5) | 0.33 ± 0.20 | 0.05 | 0.18 | 0.47 | 0.98 |
| | Adult | 7(100) | 3.60 ± 1.85 | 2.05 | 2.37 | 3.74 | 7.46 |
| Treg | Birth | 176(88) | 0.21 ± 0.10 | 0.05 | 0.14 | 0.26 | 0.60 |
| | Month-6 | 70(35) | 0.30 ± 0.17 | 0.05 | 0.19 | 0.34 | 1.07 |
| | Month-12 | 55(27.5) | 0.39 ± 0.24 | 0.11 | 0.24 | 0.50 | 1.66 |
| | Adult | 7(100) | 2.11 ± 0.95 | 1.30 | 1.51 | 2.28 | 4.07 |
| Th17 | Birth | 176(88) | 0.00 ± 0.01 | 0.00 | 0.00 | 0.00 | 0.06 |
| | Month-6 | 70(35) | 0.07 ± 0.06 | 0.01 | 0.03 | 0.09 | 0.43 |
| | Month-12 | 55(27.5) | 0.12 ± 0.09 | 0.02 | 0.06 | 0.15 | 0.43 |
| | Adult | 7(100) | 0.62 ± 0.30 | 0.21 | 0.44 | 0.78 | 1.10 |

Tfh: T follicular helper cell, Th2: T helper 2 cell, Th1: T helper 1 cell, Treg: T regulatory cell, Th17: T helper 17 cell, SD: standard deviation, Min: minimum, Max: maximum, Q: quantile.

**Supplemental Table 6:** Average marginal effects for age-specific models associating maternal PFAS concentrations ($log_2$) and infant CD4+ T-cells.

| Cell Type | PFAS | Infant Age | Unadjusted | | Adjusted | |
|---|---|---|---|---|---|---|
| | | | AME (95% CI) | P-value | AME (95% CI) | P-value |
| Tfh | PFOS | Birth | 0.06 (-0.03, 0.14) | 0.195 | 0.08 (-0.01, 0.16) | 0.082 |
| | | Month-6 | -0.07 (-0.18, 0.04) | 0.206 | -0.05 (-0.16, 0.06) | 0.336 |
| | | Month-12 | -0.18 (-0.32, -0.05) | 0.007 | -0.17 (-0.30, -0.04) | 0.013 |
| | PFOA | Birth | 0.00 (-0.14, 0.14) | 0.947 | 0.04 (-0.11, 0.19) | 0.626 |
| | | Month-6 | -0.04 (-0.25, 0.16) | 0.688 | 0.02 (-0.20, 0.23) | 0.891 |
| | | Month-12 | -0.20 (-0.39, -0.01) | 0.035 | -0.17 (-0.36, 0.03) | 0.089 |
| | PFNA | Birth | -0.02 (-0.18, 0.15) | 0.842 | 0.00 (-0.16, 0.17) | 0.971 |
| | | Month-6 | -0.12 (-0.35, 0.11) | 0.290 | -0.11 (-0.33, 0.12) | 0.357 |
| | | Month-12 | -0.25 (-0.48, -0.01) | 0.037 | -0.25 (-0.48, -0.02) | 0.034 |
| | PFHXS | Birth | -0.01 (-0.13, 0.10) | 0.829 | 0.01 (-0.11, 0.13) | 0.879 |
| | | Month-6 | -0.08 (-0.24, 0.08) | 0.327 | -0.05 (-0.21, 0.11) | 0.530 |
| | | Month-12 | -0.23 (-0.43, -0.03) | 0.021 | -0.19 (-0.39, 0.01) | 0.062 |
| Th2 | PFOS | Birth | -0.01 (-0.06, 0.04) | 0.829 | 0.01 (-0.04, 0.06) | 0.720 |
| | | Month-6 | 0.10 (0.03, 0.16) | 0.005 | 0.11 (0.04, 0.18) | 0.001 |
| | | Month-12 | 0.25 (0.17, 0.34) | 0.000 | 0.27 (0.18, 0.35) | 0.000 |
| | PFOA | Birth | -0.01 (-0.09, 0.08) | 0.900 | 0.05 (-0.04, 0.14) | 0.248 |
| | | Month-6 | 0.18 (0.05, 0.31) | 0.008 | 0.25 (0.12, 0.38) | 0.000 |
| | | Month-12 | 0.25 (0.13, 0.36) | 0.000 | 0.30 (0.18, 0.41) | 0.000 |
| | PFNA | Birth | -0.01 (-0.1, 0.09) | 0.882 | 0.00 (-0.09, 0.09) | 0.991 |
| | | Month-6 | 0.31 (0.18, 0.45) | 0.000 | 0.33 (0.20, 0.47) | 0.000 |
| | | Month-12 | 0.46 (0.32, 0.59) | 0.000 | 0.46 (0.32, 0.60) | 0.000 |
| | PFHXS | Birth | 0.00 (-0.07, 0.07) | 0.972 | 0.02 (-0.06, 0.09) | 0.658 |
| | | Month-6 | 0.04 (-0.06, 0.14) | 0.477 | 0.06 (-0.05, 0.16) | 0.287 |
| | | Month-12 | 0.11 (-0.01, 0.24) | 0.080 | 0.14 (0.01, 0.27) | 0.032 |
| Th1 | PFOS | Birth | -0.01 (-0.04, 0.02) | 0.599 | -0.01 (-0.04, 0.02) | 0.561 |
| | | Month-6 | -0.04 (-0.08, 0.00) | 0.067 | -0.04 (-0.08, 0.00) | 0.069 |
| | | Month-12 | 0.02 (-0.03, 0.07) | 0.435 | 0.02 (-0.03, 0.07) | 0.439 |
| | PFOA | Birth | -0.01 (-0.06, 0.04) | 0.630 | -0.01 (-0.06, 0.05) | 0.768 |
| | | Month-6 | 0.00 (-0.08, 0.08) | 0.988 | 0.01 (-0.07, 0.09) | 0.840 |
| | | Month-12 | 0.12 (0.05, 0.19) | 0.001 | 0.12 (0.05, 0.19) | 0.001 |
| | | Birth | -0.01 (-0.07, 0.05) | 0.701 | -0.01 (-0.07, 0.05) | 0.764 |

| | | | | | | |
|---|---|---|---|---|---|---|
| | PFNA | Month-6 | 0.00 (-0.09, 0.08) | 0.914 | 0.00 (-0.09, 0.09) | 0.959 |
| | | Month-12 | 0.11 (0.02, 0.20) | 0.014 | 0.11 (0.02, 0.20) | 0.015 |
| | PFHXS | Birth | 0.00 (-0.04, 0.04) | 0.925 | 0.00 (-0.04, 0.04) | 0.958 |
| | | Month-6 | -0.04 (-0.10, 0.02) | 0.234 | -0.03 (-0.10, 0.03) | 0.270 |
| | | Month-12 | 0.05 (-0.03, 0.12) | 0.245 | 0.05 (-0.03, 0.13) | 0.203 |
| Treg | PFOS | Birth | 0.00 (-0.04, 0.04) | 0.982 | 0.00 (-0.04, 0.04) | 0.938 |
| | | Month-6 | 0.00 (-0.06, 0.05) | 0.966 | 0.00 (-0.06, 0.06) | 0.968 |
| | | Month-12 | 0.04 (-0.03, 0.11) | 0.306 | 0.04 (-0.03, 0.11) | 0.312 |
| | PFOA | Birth | -0.01 (-0.08, 0.06) | 0.723 | -0.01 (-0.08, 0.06) | 0.784 |
| | | Month-6 | 0.03 (-0.08, 0.14) | 0.576 | 0.04 (-0.07, 0.15) | 0.517 |
| | | Month-12 | 0.07 (-0.02, 0.17) | 0.137 | 0.07 (-0.02, 0.17) | 0.140 |
| | PFNA | Birth | -0.05 (-0.13, 0.03) | 0.210 | -0.05 (-0.13, 0.03) | 0.237 |
| | | Month-6 | 0.04 (-0.08, 0.16) | 0.502 | 0.04 (-0.08, 0.16) | 0.486 |
| | | Month-12 | 0.06 (-0.06, 0.18) | 0.333 | 0.06 (-0.06, 0.18) | 0.328 |
| | PFHXS | Birth | 0.00 (-0.05, 0.06) | 0.932 | 0.00 (-0.05, 0.06) | 0.881 |
| | | Month-6 | 0.06 (-0.02, 0.14) | 0.135 | 0.06 (-0.02, 0.15) | 0.121 |
| | | Month-12 | 0.11 (0.01, 0.21) | 0.039 | 0.11 (0.01, 0.22) | 0.031 |
| Th17 | PFOS | Birth | 0.00 (-0.01, 0.01) | 0.935 | 0.00 (-0.01, 0.01) | 0.973 |
| | | Month-6 | -0.01 (-0.03, 0.01) | 0.414 | -0.01 (-0.03, 0.01) | 0.488 |
| | | Month-12 | -0.01 (-0.04, 0.01) | 0.251 | -0.01 (-0.04, 0.01) | 0.288 |
| | PFOA | Birth | 0.00 (-0.02, 0.02) | 0.977 | 0.00 (-0.02, 0.03) | 0.712 |
| | | Month-6 | 0.00 (-0.04, 0.04) | 0.988 | 0.01 (-0.03, 0.04) | 0.682 |
| | | Month-12 | -0.01 (-0.04, 0.02) | 0.639 | 0.00 (-0.04, 0.03) | 0.841 |
| | PFNA | Birth | 0.00 (-0.02, 0.03) | 0.800 | 0.01 (-0.02, 0.03) | 0.670 |
| | | Month-6 | -0.01 (-0.05, 0.03) | 0.697 | 0.00 (-0.04, 0.03) | 0.837 |
| | | Month-12 | 0.00 (-0.04, 0.04) | 0.865 | 0.00 (-0.04, 0.04) | 0.925 |
| | PFHXS | Birth | 0.00 (-0.02, 0.02) | 0.958 | 0.00 (-0.02, 0.02) | 0.980 |
| | | Month-6 | -0.01 (-0.03, 0.02) | 0.592 | -0.01 (-0.03, 0.02) | 0.612 |
| | | Month-12 | 0.01 (-0.02, 0.05) | 0.432 | 0.02 (-0.02, 0.05) | 0.350 |

Tfh: T follicular helper cell, Th2: T helper 2 cell, Th1: T helper 1 cell, Treg: T regulatory cell, Th17: T helper 17 cell, PFOS: perfluorooctanesulfonic acid, PFOA: perfluorooctanoic acid, PFNA: perfluorononanoic acid, PFHxS: perfluorohexane sulfonic acid, AME: average marginal effect, CI: confidence intervals. Unadjusted and adjusted Linear Mixed-Effects (LMER).

**Supplemental Table 7:** Unadjusted and adjusted interaction P-values between PFAS concentrations ($log_2$) and infant age for age-specific models associating maternal PFAS concentrations and infant CD4+ T-cells.

| Cell Type | PFAS | Unadjusted P-value | Adjusted P-value |
|---|---|---|---|
| Tfh | PFOS | 0.002 | 0.002 |
| | PFOA | 0.138 | 0.112 |
| | PFNA | 0.168 | 0.120 |
| | PFHXS | 0.115 | 0.167 |
| Th2 | PFOS | 0.000 | 0.000 |
| | PFOA | 0.000 | 0.000 |
| | PFNA | 0.000 | 0.000 |
| | PFHXS | 0.271 | 0.215 |
| Th1 | PFOS | 0.174 | 0.176 |
| | PFOA | 0.007 | 0.008 |
| | PFNA | 0.047 | 0.054 |
| | PFHXS | 0.226 | 0.203 |
| Treg | PFOS | 0.628 | 0.620 |
| | PFOA | 0.334 | 0.333 |
| | PFNA | 0.211 | 0.221 |
| | PFHXS | 0.150 | 0.129 |
| Th17 | PFOS | 0.591 | 0.631 |
| | PFOA | 0.925 | 0.884 |
| | PFNA | 0.884 | 0.897 |
| | PFHXS | 0.623 | 0.549 |

Tfh: T follicular helper cell, Th2: T helper 2 cell, Th1: T helper 1 cell, Treg: T regulatory cell, Th17: T helper 17 cell, PFOS: perfluorooctanesulfonic acid, PFOA: perfluorooctanoic acid, PFNA: perfluorononanoic acid, PFHxS: perfluorohexane sulfonic acid, df: degrees of freedom.

**Supplemental Table 8**: Unadjusted and adjusted pairwise comparison of the average marginal effects for age-specific models associating maternal PFAS concentrations and infant CD4+ T-cells.

| Cell Type | PFAS | Comparison | Unadjusted | | Adjusted | |
|---|---|---|---|---|---|---|
| | | | Contrast (95% CI) | P-value | Contrast (95% CI) | P-value |
| Tfh | PFOS | (Birth)-(Month-6) | 0.126 (0.01, 0.24) | 0.037 | 0.128 (0.01, 0.25) | 0.033 |
| | | (Birth)-(Month-12) | 0.240 (0.10, 0.38) | 0.001 | 0.245 (0.10, 0.39) | 0.001 |
| | | (Month-6)-(Month-12) | 0.114 (-0.04, 0.27) | 0.143 | 0.117 (-0.04, 0.27) | 0.134 |
| | PFOA | (Birth)-(Month-6) | 0.038 (-0.19, 0.26) | 0.739 | 0.022 (-0.20, 0.24) | 0.846 |
| | | (Birth)-(Month-12) | 0.195 (0.00, 0.39) | 0.050 | 0.203 (0.01, 0.40) | 0.041 |
| | | (Month-6)-(Month-12) | 0.157 (-0.09, 0.41) | 0.216 | 0.181 (-0.07, 0.43) | 0.155 |
| | PFNA | (Birth)-(Month-6) | 0.107 (-0.14, 0.35) | 0.385 | 0.110 (-0.13, 0.35) | 0.373 |
| | | (Birth)-(Month-12) | 0.228 (-0.01, 0.47) | 0.062 | 0.250 (0.01, 0.49) | 0.041 |
| | | (Month-6)-(Month-12) | 0.121 (-0.15, 0.39) | 0.383 | 0.140 (-0.13, 0.41) | 0.316 |
| | PFHXS | (Birth)-(Month-6) | 0.067 (-0.11, 0.24) | 0.454 | 0.060 (-0.11, 0.23) | 0.496 |
| | | (Birth)-(Month-12) | 0.217 (0.01, 0.42) | 0.039 | 0.197 (-0.01, 0.40) | 0.059 |
| | | (Month-6)-(Month-12) | 0.151 (-0.08, 0.38) | 0.192 | 0.137 (-0.09, 0.36) | 0.234 |
| Th2 | PFOS | (Birth)-(Month-6) | -0.101 (-0.18, -0.02) | 0.012 | -0.101 (-0.18, -0.02) | 0.013 |
| | | (Birth)-(Month-12) | -0.258 (-0.35, -0.16) | 0.000 | -0.257 (-0.35, -0.16) | 0.000 |
| | | (Month-6)-(Month-12) | -0.157 (-0.26, -0.05) | 0.003 | -0.156 (-0.26, -0.05) | 0.003 |
| | PFOA | (Birth)-(Month-6) | -0.181 (-0.33, -0.03) | 0.017 | -0.196 (-0.34, -0.05) | 0.010 |
| | | (Birth)-(Month-12) | -0.254 (-0.39, -0.12) | 0.000 | -0.247 (-0.38, -0.11) | 0.000 |
| | | (Month-6)-(Month-12) | -0.072 (-0.24, 0.10) | 0.397 | -0.050 (-0.22, 0.12) | 0.555 |
| | PFNA | (Birth)-(Month-6) | -0.322 (-0.48, -0.16) | 0.000 | -0.333 (-0.49, -0.17) | 0.000 |
| | | (Birth)-(Month-12) | -0.463 (-0.62, -0.31) | 0.000 | -0.460 (-0.62, -0.30) | 0.000 |
| | | (Month-6)-(Month-12) | -0.141 (-0.32, 0.04) | 0.129 | -0.128 (-0.31, 0.06) | 0.173 |
| | PFHXS | (Birth)-(Month-6) | -0.038 (-0.16, 0.08) | 0.530 | -0.039 (-0.16, 0.08) | 0.518 |
| | | (Birth)-(Month-12) | -0.116 (-0.26, 0.03) | 0.109 | -0.126 (-0.27, 0.02) | 0.082 |
| | | (Month-6)-(Month-12) | -0.078 (-0.23, 0.08) | 0.331 | -0.087 (-0.24, 0.07) | 0.278 |
| Th1 | PFOS | (Birth)-(Month-6) | 0.031 (-0.02, 0.08) | 0.225 | 0.030 (-0.02, 0.08) | 0.240 |
| | | (Birth)-(Month-12) | -0.029 (-0.09, 0.03) | 0.328 | -0.030 (-0.09, 0.03) | 0.311 |
| | | (Month-6)-(Month-12) | -0.06 (-0.12, 0.00) | 0.068 | -0.060 (-0.12, 0.00) | 0.067 |
| | PFOA | (Birth)-(Month-6) | -0.012 (-0.1, 0.08) | 0.797 | -0.017 (-0.11, 0.07) | 0.719 |
| | | (Birth)-(Month-12) | -0.127 (-0.21, -0.05) | 0.002 | -0.127 (-0.21, -0.05) | 0.002 |
| | | (Month-6)-(Month-12) | -0.116 (-0.22, -0.01) | 0.026 | -0.11 (-0.21, -0.01) | 0.035 |
| | | (Birth)-(Month-6) | -0.007 (-0.11, 0.09) | 0.893 | -0.007 (-0.11, 0.09) | 0.893 |

| | | | | | | |
|---|---|---|---|---|---|---|
| | PFNA | (Birth)-(Month-12) | -0.121 (-0.22, -0.02) | 0.018 | -0.119 (-0.22, -0.02) | 0.021 |
| | | (Month-6)-(Month-12) | -0.114 (-0.23, 0.00) | 0.054 | -0.112 (-0.23, 0.00) | 0.059 |
| | PFHXS | (Birth)-(Month-6) | 0.034 (-0.04, 0.11) | 0.338 | 0.033 (-0.04, 0.10) | 0.356 |
| | | (Birth)-(Month-12) | -0.047 (-0.13, 0.04) | 0.271 | -0.052 (-0.14, 0.03) | 0.231 |
| | | (Month-6)-(Month-12) | -0.082 (-0.18, 0.01) | 0.086 | -0.085 (-0.18, 0.01) | 0.075 |
| Treg | PFOS | (Birth)-(Month-6) | 0.001 (-0.07, 0.07) | 0.983 | -0.001 (-0.07, 0.07) | 0.988 |
| | | (Birth)-(Month-12) | -0.038 (-0.12, 0.04) | 0.362 | -0.039 (-0.12, 0.04) | 0.350 |
| | | (Month-6)-(Month-12) | -0.038 (-0.13, 0.05) | 0.395 | -0.038 (-0.13, 0.05) | 0.399 |
| | PFOA | (Birth)-(Month-6) | -0.043 (-0.17, 0.08) | 0.499 | -0.047 (-0.17, 0.08) | 0.462 |
| | | (Birth)-(Month-12) | -0.084 (-0.20, 0.03) | 0.145 | -0.084 (-0.20, 0.03) | 0.148 |
| | | (Month-6)-(Month-12) | -0.042 (-0.18, 0.10) | 0.561 | -0.037 (-0.18, 0.10) | 0.607 |
| | PFNA | (Birth)-(Month-6) | -0.091 (-0.23, 0.05) | 0.196 | -0.091 (-0.23, 0.05) | 0.201 |
| | | (Birth)-(Month-12) | -0.110 (-0.25, 0.03) | 0.122 | -0.108 (-0.25, 0.03) | 0.128 |
| | | (Month-6)-(Month-12) | -0.018 (-0.18, 0.14) | 0.824 | -0.018 (-0.18, 0.14) | 0.832 |
| | PFHXS | (Birth)-(Month-6) | -0.059 (-0.15, 0.04) | 0.230 | -0.060 (-0.16, 0.04) | 0.219 |
| | | (Birth)-(Month-12) | -0.105 (-0.22, 0.01) | 0.072 | -0.110 (-0.23, 0.00) | 0.060 |
| | | (Month-6)-(Month-12) | -0.046 (-0.17, 0.08) | 0.473 | -0.050 (-0.18, 0.08) | 0.442 |
| Th17 | PFOS | (Birth)-(Month-6) | 0.007 (-0.02, 0.03) | 0.536 | 0.006 (-0.02, 0.03) | 0.583 |
| | | (Birth)-(Month-12) | 0.013 (-0.01, 0.04) | 0.335 | 0.013 (-0.01, 0.04) | 0.360 |
| | | (Month-6)-(Month-12) | 0.006 (-0.02, 0.04) | 0.685 | 0.006 (-0.02, 0.04) | 0.678 |
| | PFOA | (Birth)-(Month-6) | 0.000 (-0.04, 0.04) | 0.997 | -0.003 (-0.04, 0.04) | 0.882 |
| | | (Birth)-(Month-12) | 0.007 (-0.03, 0.04) | 0.708 | 0.008 (-0.03, 0.05) | 0.685 |
| | | (Month-6)-(Month-12) | 0.007 (-0.04, 0.05) | 0.760 | 0.011 (-0.04, 0.06) | 0.646 |
| | PFNA | (Birth)-(Month-6) | 0.011 (-0.03, 0.06) | 0.635 | 0.010 (-0.04, 0.06) | 0.674 |
| | | (Birth)-(Month-12) | 0.007 (-0.04, 0.05) | 0.771 | 0.008 (-0.04, 0.05) | 0.745 |
| | | (Month-6)-(Month-12) | -0.004 (-0.06, 0.05) | 0.875 | -0.002 (-0.06, 0.05) | 0.935 |
| | PFHXS | (Birth)-(Month-6) | 0.007 (-0.03, 0.04) | 0.676 | 0.007 (-0.03, 0.04) | 0.679 |
| | | (Birth)-(Month-12) | -0.014 (-0.05, 0.02) | 0.468 | -0.017 (-0.06, 0.02) | 0.392 |
| | | (Month-6)-(Month-12) | -0.021 (-0.06, 0.02) | 0.333 | -0.023 (-0.07, 0.02) | 0.279 |

Tfh: T follicular helper cell, Th2: T helper 2 cell, Th1: T helper 1 cell, Treg: T regulatory cell, Th17: T helper 17 cell, PFOS: perfluorooctanesulfonic acid, PFOA: perfluorooctanoic acid, PFNA: perfluorononanoic acid, PFHxS: perfluorohexane sulfonic acid, CI: confidence intervals.

**Supplemental Table 9:** Unadjusted and adjusted predicted marginal means of infant CD4+ T-cells by categorical maternal PFAS for differences-over-time models.

| Cell Type | PFAS | Visit | PFAS Value | Unadjusted | | | | Adjusted | | | |
|---|---|---|---|---|---|---|---|---|---|---|---|
| | | | | LSMEAN (SE) (95%CI) | df | t-ratio | P-value | LSMEAN (SE) (95%CI) | df | t-ratio | P-value |
| Tfh | PFOS | Birth | Below | 0.344 (0.03) (0.28, 0.41) | 269.6 | 10.9 | 0.000 | 0.329 (0.03) (0.27, 0.39) | 263.4 | 10.3 | 0.000 |
| | | | Above | 0.343 (0.03) (0.28, 0.41) | 280.1 | 10.4 | 0.000 | 0.360 (0.03) (0.29, 0.43) | 272.6 | 10.7 | 0.000 |
| | | Month-6 | Below | 0.618 (0.05) (0.52, 0.72) | 270.6 | 11.8 | 0.000 | 0.603 (0.05) (0.50, 0.71) | 271.5 | 11.5 | 0.000 |
| | | | Above | 0.555 (0.05) (0.46, 0.64) | 284.4 | 12.1 | 0.000 | 0.564 (0.05) (0.47, 0.65) | 283.9 | 12.2 | 0.000 |
| | | Month-12 | Below | 1.128 (0.06) (1.02, 1.24) | 250.1 | 20.0 | 0.000 | 1.106 (0.06) (0.99, 1.22) | 250.8 | 19.5 | 0.000 |
| | | | Above | 0.859 (0.05) (0.76, 0.96) | 269.9 | 16.5 | 0.000 | 0.864 (0.05) (0.76, 0.97) | 270.6 | 16.4 | 0.000 |
| | PFOA | Birth | Below | 0.367 (0.03) (0.30, 0.43) | 273.2 | 11.2 | 0.000 | 0.359 (0.03) (0.29, 0.43) | 265.6 | 10.8 | 0.000 |
| | | | Above | 0.316 (0.03) (0.25, 0.38) | 275.4 | 9.6 | 0.000 | 0.326 (0.03) (0.26, 0.39) | 261.0 | 9.3 | 0.000 |
| | | Month-6 | Below | 0.604 (0.05) (0.50, 0.70) | 280.0 | 12.0 | 0.000 | 0.588 (0.05) (0.49, 0.69) | 280.7 | 11.6 | 0.000 |
| | | | Above | 0.562 (0.05) (0.47, 0.66) | 276.8 | 11.6 | 0.000 | 0.573 (0.05) (0.47, 0.67) | 282.4 | 11.5 | 0.000 |
| | | Month-12 | Below | 1.038 (0.06) (0.92, 1.15) | 249.0 | 18.1 | 0.000 | 1.022 (0.06) (0.91, 1.14) | 252.1 | 17.7 | 0.000 |
| | | | Above | 0.936 (0.05) (0.83, 1.04) | 270.0 | 17.7 | 0.000 | 0.937 (0.05) (0.83, 1.04) | 276.0 | 17.2 | 0.000 |
| | PFNA | Birth | Below | 0.339 (0.03) (0.28, 0.40) | 272.8 | 10.7 | 0.000 | 0.331 (0.03) (0.27, 0.39) | 270.0 | 10.5 | 0.000 |
| | | | Above | 0.345 (0.03) (0.28, 0.41) | 276.0 | 10.2 | 0.000 | 0.358 (0.03) (0.29, 0.42) | 269.4 | 10.5 | 0.000 |
| | | Month-6 | Below | 0.580 (0.05) (0.49, 0.67) | 278.9 | 12.3 | 0.000 | 0.568 (0.05) (0.48, 0.66) | 276.0 | 12.1 | 0.000 |
| | | | Above | 0.590 (0.05) (0.49, 0.69) | 277.4 | 11.5 | 0.000 | 0.601 (0.05) (0.50, 0.70) | 276.7 | 11.6 | 0.000 |
| | | Month-12 | Below | 1.057 (0.05) (0.95, 1.16) | 257.7 | 19.8 | 0.000 | 1.043 (0.05) (0.94, 1.15) | 257.7 | 19.7 | 0.000 |
| | | | Above | 0.900 (0.06) (0.79, 1.01) | 263.5 | 15.9 | 0.000 | 0.902 (0.06) (0.79, 1.01) | 266.3 | 15.9 | 0.000 |
| | PFHXS | Birth | Below | 0.337 (0.03) (0.27, 0.40) | 273.9 | 9.9 | 0.000 | 0.335 (0.03) (0.27, 0.40) | 270.2 | 9.9 | 0.000 |
| | | | Above | 0.345 (0.03) (0.28, 0.41) | 274.9 | 10.8 | 0.000 | 0.351 (0.03) (0.29, 0.41) | 270.1 | 10.9 | 0.000 |
| | | Month-6 | Below | 0.591 (0.05) (0.49, 0.69) | 277.3 | 11.2 | 0.000 | 0.581 (0.05) (0.48, 0.68) | 275.8 | 11.1 | 0.000 |
| | | | Above | 0.577 (0.05) (0.48, 0.67) | 279.1 | 12.3 | 0.000 | 0.584 (0.05) (0.49, 0.68) | 277.8 | 12.4 | 0.000 |
| | | Month-12 | Below | 1.011 (0.06) (0.89, 1.13) | 255.2 | 17.2 | 0.000 | 0.994 (0.06) (0.88, 1.11) | 255.3 | 17.0 | 0.000 |
| | | | Above | 0.962 (0.05) (0.86, 1.06) | 264.7 | 18.5 | 0.000 | 0.965 (0.05) (0.86, 1.07) | 266.0 | 18.5 | 0.000 |
| Th2 | PFOS | Birth | Below | 0.016 (0.02) (-0.02, 0.05) | 289.7 | 0.8 | 0.847 | 0.008 (0.02) (-0.03, 0.05) | 283.6 | 0.4 | 1.000 |
| | | | Above | 0.014 (0.02) (-0.03, 0.05) | 292.2 | 0.7 | 0.986 | 0.030 (0.02) (-0.01, 0.07) | 285.9 | 1.4 | 0.319 |
| | | Month-6 | Below | 0.146 (0.03) (0.08, 0.21) | 288.1 | 4.3 | 0.000 | 0.135 (0.03) (0.07, 0.20) | 286.7 | 3.9 | 0.000 |
| | | | Above | 0.193 (0.03) (0.13, 0.25) | 291.8 | 6.5 | 0.000 | 0.210 (0.03) (0.15, 0.27) | 289.4 | 7.0 | 0.000 |
| | | Month-12 | Below | 0.329 (0.04) (0.26, 0.40) | 281.2 | 8.8 | 0.000 | 0.313 (0.04) (0.24, 0.39) | 280.1 | 8.3 | 0.000 |
| | | | Above | 0.427 (0.03) (0.36, 0.49) | 287.2 | 12.5 | 0.000 | 0.446 (0.03) (0.38, 0.51) | 286.0 | 12.8 | 0.000 |

| | | | | | | | | | | | |
|---|---|---|---|---|---|---|---|---|---|---|---|
| | PFOA | Birth | Below | 0.017 (0.02) (-0.02, 0.06) | 290.9 | 0.9 | 0.779 | 0.004 (0.02) (-0.04, 0.04) | 284.7 | 0.2 | 1.000 |
| | | | Above | 0.013 (0.02) (-0.03, 0.05) | 291.4 | 0.7 | 1.000 | 0.036 (0.02) (-0.01, 0.08) | 280.0 | 1.7 | 0.175 |
| | | Month-6 | Below | 0.133 (0.03) (0.07, 0.20) | 291.2 | 4.2 | 0.000 | 0.117 (0.03) (0.05, 0.18) | 289.4 | 3.7 | 0.001 |
| | | | Above | 0.210 (0.03) (0.15, 0.27) | 289.7 | 6.8 | 0.000 | 0.236 (0.03) (0.17, 0.30) | 290.0 | 7.4 | 0.000 |
| | | Month-12 | Below | 0.319 (0.04) (0.25, 0.39) | 281.8 | 8.6 | 0.000 | 0.298 (0.04) (0.22, 0.37) | 282.8 | 7.9 | 0.000 |
| | | | Above | 0.436 (0.03) (0.37, 0.50) | 287.6 | 12.8 | 0.000 | 0.462 (0.03) (0.39, 0.53) | 288.8 | 13.3 | 0.000 |
| | PFNA | Birth | Below | 0.017 (0.02) (-0.02, 0.05) | 290.8 | 0.9 | 0.771 | 0.013 (0.02) (-0.02, 0.05) | 286.6 | 0.7 | 0.989 |
| | | | Above | 0.013 (0.02) (-0.03, 0.05) | 291.4 | 0.6 | 1.000 | 0.025 (0.02) (-0.02, 0.07) | 285.2 | 1.2 | 0.454 |
| | | Month-6 | Below | 0.138 (0.03) (0.08, 0.20) | 290.3 | 4.6 | 0.000 | 0.130 (0.03) (0.07, 0.19) | 286.8 | 4.3 | 0.000 |
| | | | Above | 0.213 (0.03) (0.15, 0.28) | 290.2 | 6.5 | 0.000 | 0.230 (0.03) (0.16, 0.30) | 287.5 | 6.9 | 0.000 |
| | | Month-12 | Below | 0.309 (0.03) (0.24, 0.38) | 283.5 | 9.0 | 0.000 | 0.305 (0.03) (0.24, 0.37) | 282.0 | 8.9 | 0.000 |
| | | | Above | 0.464 (0.04) (0.39, 0.53) | 286.0 | 12.8 | 0.000 | 0.475 (0.04) (0.40, 0.55) | 285.6 | 13.0 | 0.000 |
| | PFHXS | Birth | Below | 0.016 (0.02) (-0.02, 0.06) | 291.1 | 0.8 | 0.879 | 0.014 (0.02) (-0.03, 0.06) | 286.2 | 0.7 | 1.000 |
| | | | Above | 0.015 (0.02) (-0.02, 0.05) | 291.4 | 0.7 | 0.910 | 0.022 (0.02) (-0.02, 0.06) | 286.2 | 1.1 | 0.543 |
| | | Month-6 | Below | 0.150 (0.03) (0.08, 0.22) | 290.4 | 4.4 | 0.000 | 0.148 (0.03) (0.08, 0.22) | 287.3 | 4.4 | 0.000 |
| | | | Above | 0.191 (0.03) (0.13, 0.25) | 290.4 | 6.3 | 0.000 | 0.198 (0.03) (0.14, 0.26) | 287.2 | 6.5 | 0.000 |
| | | Month-12 | Below | 0.346 (0.04) (0.27, 0.42) | 283.8 | 9.0 | 0.000 | 0.340 (0.04) (0.26, 0.42) | 282.1 | 8.8 | 0.000 |
| | | | Above | 0.410 (0.03) (0.34, 0.48) | 286.1 | 12.2 | 0.000 | 0.419 (0.03) (0.35, 0.49) | 284.7 | 12.3 | 0.000 |
| Th1 | PFOS | Birth | Below | 0.045 (0.01) (0.02, 0.07) | 290.1 | 3.8 | 0.000 | 0.044 (0.01) (0.02, 0.07) | 282.4 | 3.7 | 0.001 |
| | | | Above | 0.038 (0.01) (0.01, 0.06) | 292.4 | 3.1 | 0.004 | 0.038 (0.01) (0.01, 0.06) | 285.1 | 3.0 | 0.005 |
| | | Month-6 | Below | 0.200 (0.02) (0.16, 0.24) | 288.6 | 9.8 | 0.000 | 0.198 (0.02) (0.16, 0.24) | 285.6 | 9.6 | 0.000 |
| | | | Above | 0.172 (0.02) (0.14, 0.21) | 292.0 | 9.7 | 0.000 | 0.170 (0.02) (0.13, 0.21) | 289.0 | 9.4 | 0.000 |
| | | Month-12 | Below | 0.314 (0.02) (0.27, 0.36) | 282.2 | 14.0 | 0.000 | 0.311 (0.02) (0.27, 0.36) | 277.8 | 13.7 | 0.000 |
| | | | Above | 0.341 (0.02) (0.30, 0.38) | 287.8 | 16.7 | 0.000 | 0.339 (0.02) (0.30, 0.38) | 284.8 | 16.2 | 0.000 |
| | PFOA | Birth | Below | 0.046 (0.01) (0.02, 0.07) | 291.0 | 3.9 | 0.000 | 0.047 (0.01) (0.02, 0.07) | 282.8 | 3.8 | 0.000 |
| | | | Above | 0.037 (0.01) (0.01, 0.06) | 291.5 | 3.0 | 0.005 | 0.036 (0.01) (0.01, 0.06) | 277.9 | 2.8 | 0.011 |
| | | Month-6 | Below | 0.189 (0.02) (0.15, 0.23) | 291.3 | 9.8 | 0.000 | 0.187 (0.02) (0.15, 0.23) | 288.5 | 9.5 | 0.000 |
| | | | Above | 0.179 (0.02) (0.14, 0.22) | 289.8 | 9.6 | 0.000 | 0.178 (0.02) (0.14, 0.22) | 289.2 | 9.2 | 0.000 |
| | | Month-12 | Below | 0.306 (0.02) (0.26, 0.35) | 282.2 | 13.7 | 0.000 | 0.305 (0.02) (0.26, 0.35) | 279.0 | 13.4 | 0.000 |
| | | | Above | 0.347 (0.02) (0.31, 0.39) | 287.8 | 17.0 | 0.000 | 0.344 (0.02) (0.30, 0.39) | 287.4 | 16.3 | 0.000 |
| | PFNA | Birth | Below | 0.046 (0.01) (0.02, 0.07) | 291.1 | 4.0 | 0.000 | 0.046 (0.01) (0.02, 0.07) | 285.7 | 4.0 | 0.000 |
| | | | Above | 0.036 (0.01) (0.01, 0.06) | 291.7 | 2.9 | 0.007 | 0.036 (0.01) (0.01, 0.06) | 284.3 | 2.9 | 0.009 |
| | | Month-6 | Below | 0.185 (0.02) (0.15, 0.22) | 290.6 | 10.2 | 0.000 | 0.183 (0.02) (0.15, 0.22) | 286.0 | 10.0 | 0.000 |
| | | | Above | 0.182 (0.02) (0.14, 0.22) | 290.5 | 9.3 | 0.000 | 0.181 (0.02) (0.14, 0.22) | 286.7 | 9.0 | 0.000 |

| | | | | | | | | | | | |
|---|---|---|---|---|---|---|---|---|---|---|---|
| | | Month-12 | Below | 0.295 (0.02) (0.25, 0.34) | 284.3 | 14.3 | 0.000 | 0.293 (0.02) (0.25, 0.33) | 280.2 | 14.1 | 0.000 |
| | | | Above | 0.366 (0.02) (0.32, 0.41) | 286.7 | 16.8 | 0.000 | 0.364 (0.02) (0.32, 0.41) | 284.3 | 16.5 | 0.000 |
| | PFHXS | Birth | Below | 0.043 (0.01) (0.02, 0.07) | 291.0 | 3.5 | 0.001 | 0.044 (0.01) (0.02, 0.07) | 285.2 | 3.5 | 0.001 |
| | | | Above | 0.040 (0.01) (0.02, 0.06) | 291.3 | 3.5 | 0.001 | 0.040 (0.01) (0.02, 0.06) | 285.2 | 3.3 | 0.002 |
| | | Month-6 | Below | 0.193 (0.02) (0.15, 0.23) | 290.4 | 9.6 | 0.000 | 0.191 (0.02) (0.15, 0.23) | 286.4 | 9.4 | 0.000 |
| | | | Above | 0.177 (0.02) (0.14, 0.21) | 290.4 | 9.9 | 0.000 | 0.176 (0.02) (0.14, 0.21) | 286.4 | 9.7 | 0.000 |
| | | Month-12 | Below | 0.305 (0.02) (0.26, 0.35) | 283.7 | 13.4 | 0.000 | 0.302 (0.02) (0.26, 0.35) | 280.0 | 13.2 | 0.000 |
| | | | Above | 0.346 (0.02) (0.31, 0.39) | 286.0 | 17.3 | 0.000 | 0.345 (0.02) (0.31, 0.38) | 283.2 | 17.0 | 0.000 |
| Treg | PFOS | Birth | Below | 0.204 (0.02) (0.17, 0.23) | 293.5 | 13.1 | 0.000 | 0.203 (0.02) (0.17, 0.23) | 286.8 | 12.7 | 0.000 |
| | | | Above | 0.211 (0.02) (0.18, 0.24) | 294.2 | 12.9 | 0.000 | 0.212 (0.02) (0.18, 0.24) | 287.9 | 12.5 | 0.000 |
| | | Month-6 | Below | 0.324 (0.03) (0.27, 0.38) | 292.8 | 11.8 | 0.000 | 0.322 (0.03) (0.27, 0.38) | 289.3 | 11.6 | 0.000 |
| | | | Above | 0.288 (0.02) (0.24, 0.34) | 294.0 | 12.2 | 0.000 | 0.286 (0.02) (0.24, 0.33) | 290.4 | 11.8 | 0.000 |
| | | Month-12 | Below | 0.361 (0.03) (0.30, 0.42) | 290.6 | 12.0 | 0.000 | 0.359 (0.03) (0.30, 0.42) | 286.1 | 11.7 | 0.000 |
| | | | Above | 0.418 (0.03) (0.36, 0.47) | 292.4 | 15.3 | 0.000 | 0.415 (0.03) (0.36, 0.47) | 288.9 | 14.8 | 0.000 |
| | PFOA | Birth | Below | 0.213 (0.02) (0.18, 0.24) | 293.7 | 13.4 | 0.000 | 0.214 (0.02) (0.18, 0.25) | 286.7 | 13.0 | 0.000 |
| | | | Above | 0.201 (0.02) (0.17, 0.23) | 293.9 | 12.5 | 0.000 | 0.201 (0.02) (0.17, 0.23) | 282.4 | 11.7 | 0.000 |
| | | Month-6 | Below | 0.294 (0.03) (0.24, 0.34) | 293.7 | 11.4 | 0.000 | 0.291 (0.03) (0.24, 0.34) | 290.3 | 11.1 | 0.000 |
| | | | Above | 0.313 (0.03) (0.26, 0.36) | 293.2 | 12.5 | 0.000 | 0.311 (0.03) (0.26, 0.36) | 290.8 | 12.0 | 0.000 |
| | | Month-12 | Below | 0.369 (0.03) (0.31, 0.43) | 290.4 | 12.3 | 0.000 | 0.367 (0.03) (0.31, 0.43) | 286.7 | 12.0 | 0.000 |
| | | | Above | 0.411 (0.03) (0.36, 0.47) | 292.3 | 15.0 | 0.000 | 0.408 (0.03) (0.35, 0.46) | 290.2 | 14.4 | 0.000 |
| | PFNA | Birth | Below | 0.213 (0.02) (0.18, 0.24) | 293.5 | 13.9 | 0.000 | 0.214 (0.02) (0.18, 0.24) | 288.5 | 13.8 | 0.000 |
| | | | Above | 0.201 (0.02) (0.17, 0.23) | 293.7 | 12.2 | 0.000 | 0.200 (0.02) (0.17, 0.23) | 287.3 | 11.9 | 0.000 |
| | | Month-6 | Below | 0.298 (0.02) (0.25, 0.35) | 293.2 | 12.3 | 0.000 | 0.295 (0.02) (0.25, 0.34) | 288.6 | 12.1 | 0.000 |
| | | | Above | 0.311 (0.03) (0.26, 0.36) | 293.1 | 11.8 | 0.000 | 0.308 (0.03) (0.26, 0.36) | 289.2 | 11.5 | 0.000 |
| | | Month-12 | Below | 0.340 (0.03) (0.29, 0.39) | 290.4 | 12.3 | 0.000 | 0.337 (0.03) (0.28, 0.39) | 286.2 | 12.1 | 0.000 |
| | | | Above | 0.450 (0.03) (0.39, 0.51) | 291.5 | 15.4 | 0.000 | 0.448 (0.03) (0.39, 0.51) | 288.5 | 15.2 | 0.000 |
| | PFHXS | Birth | Below | 0.200 (0.02) (0.17, 0.23) | 294.0 | 12.4 | 0.000 | 0.200 (0.02) (0.17, 0.23) | 288.8 | 12.3 | 0.000 |
| | | | Above | 0.214 (0.02) (0.18, 0.24) | 294.1 | 14.0 | 0.000 | 0.214 (0.02) (0.18, 0.24) | 288.9 | 13.7 | 0.000 |
| | | Month-6 | Below | 0.266 (0.03) (0.21, 0.32) | 293.7 | 10.0 | 0.000 | 0.264 (0.03) (0.21, 0.32) | 289.7 | 9.9 | 0.000 |
| | | | Above | 0.334 (0.02) (0.29, 0.38) | 293.7 | 14.1 | 0.000 | 0.333 (0.02) (0.29, 0.38) | 289.5 | 13.9 | 0.000 |
| | | Month-12 | Below | 0.320 (0.03) (0.26, 0.38) | 291.8 | 10.6 | 0.000 | 0.317 (0.03) (0.26, 0.38) | 288.1 | 10.4 | 0.000 |
| | | | Above | 0.448 (0.03) (0.40, 0.50) | 292.4 | 16.9 | 0.000 | 0.446 (0.03) (0.39, 0.50) | 289.0 | 16.6 | 0.000 |
| Th17 | PFOS | Birth | Below | 0.002 (0.01) (-0.01, 0.01) | 293.8 | 0.3 | 1.000 | 0.002 (0.01) (-0.01, 0.01) | 287.9 | 0.3 | 1.000 |
| | | | Above | 0.001 (0.01) (-0.01, 0.01) | 294.4 | 0.3 | 1.000 | 0.003 (0.01) (-0.01, 0.01) | 288.5 | 0.5 | 1.000 |

| | | | | | | | | | | | |
|---|---|---|---|---|---|---|---|---|---|---|---|
| | | Month-6 | Below | 0.074 (0.01) (0.06, 0.09) | 293.3 | 8.1 | 0.000 | 0.072 (0.01) (0.05, 0.09) | 290.1 | 7.8 | 0.000 |
| | | | Above | 0.071 (0.01) (0.06, 0.09) | 294.2 | 9.1 | 0.000 | 0.071 (0.01) (0.06, 0.09) | 290.7 | 8.9 | 0.000 |
| | | Month-12 | Below | 0.125 (0.01) (0.11, 0.14) | 291.6 | 12.6 | 0.000 | 0.123 (0.01) (0.10, 0.14) | 288.0 | 12.2 | 0.000 |
| | | | Above | 0.123 (0.01) (0.11, 0.14) | 293.0 | 13.6 | 0.000 | 0.124 (0.01) (0.11, 0.14) | 289.8 | 13.3 | 0.000 |
| | PFOA | Birth | Below | 0.002 (0.01) (-0.01, 0.01) | 294.3 | 0.3 | 1.000 | 0.001 (0.01) (-0.01, 0.01) | 288.0 | 0.2 | 1.000 |
| | | | Above | 0.002 (0.01) (-0.01, 0.01) | 294.3 | 0.3 | 1.000 | 0.003 (0.01) (-0.01, 0.01) | 284.1 | 0.5 | 1.000 |
| | | Month-6 | Below | 0.076 (0.01) (0.06, 0.09) | 294.2 | 9.0 | 0.000 | 0.074 (0.01) (0.06, 0.09) | 290.7 | 8.6 | 0.000 |
| | | | Above | 0.068 (0.01) (0.05, 0.08) | 293.9 | 8.3 | 0.000 | 0.069 (0.01) (0.05, 0.09) | 291.0 | 8.1 | 0.000 |
| | | Month-12 | Below | 0.136 (0.01) (0.12, 0.16) | 292.3 | 13.7 | 0.000 | 0.134 (0.01) (0.11, 0.15) | 289.0 | 13.3 | 0.000 |
| | | | Above | 0.114 (0.01) (0.10, 0.13) | 293.4 | 12.6 | 0.000 | 0.114 (0.01) (0.10, 0.13) | 290.8 | 12.2 | 0.000 |
| | PFNA | Birth | Below | 0.002 (0.01) (-0.01, 0.01) | 294.0 | 0.4 | 1.000 | 0.002 (0.01) (-0.01, 0.01) | 289.4 | 0.4 | 1.000 |
| | | | Above | 0.001 (0.01) (-0.01, 0.01) | 294.2 | 0.2 | 1.000 | 0.002 (0.01) (-0.01, 0.01) | 288.4 | 0.4 | 1.000 |
| | | Month-6 | Below | 0.077 (0.01) (0.06, 0.09) | 293.8 | 9.6 | 0.000 | 0.075 (0.01) (0.06, 0.09) | 289.6 | 9.3 | 0.000 |
| | | | Above | 0.066 (0.01) (0.05, 0.08) | 293.8 | 7.6 | 0.000 | 0.067 (0.01) (0.05, 0.08) | 290.0 | 7.5 | 0.000 |
| | | Month-12 | Below | 0.121 (0.01) (0.10, 0.14) | 291.9 | 13.1 | 0.000 | 0.119 (0.01) (0.10, 0.14) | 288.3 | 12.8 | 0.000 |
| | | | Above | 0.128 (0.01) (0.11, 0.15) | 292.6 | 13.1 | 0.000 | 0.128 (0.01) (0.11, 0.15) | 289.8 | 13.1 | 0.000 |
| | PFHXS | Birth | Below | 0.001 (0.01) (-0.01, 0.01) | 294.0 | 0.3 | 1.000 | 0.002 (0.01) (-0.01, 0.01) | 289.1 | 0.4 | 1.000 |
| | | | Above | 0.002 (0.01) (-0.01, 0.01) | 294.1 | 0.3 | 1.000 | 0.002 (0.01) (-0.01, 0.01) | 289.2 | 0.4 | 1.000 |
| | | Month-6 | Below | 0.072 (0.01) (0.05, 0.09) | 293.7 | 8.0 | 0.000 | 0.071 (0.01) (0.05, 0.09) | 290.0 | 7.9 | 0.000 |
| | | | Above | 0.073 (0.01) (0.06, 0.09) | 293.7 | 9.2 | 0.000 | 0.072 (0.01) (0.06, 0.09) | 289.8 | 9.0 | 0.000 |
| | | Month-12 | Below | 0.114 (0.01) (0.09, 0.13) | 291.7 | 11.3 | 0.000 | 0.113 (0.01) (0.09, 0.13) | 288.7 | 11.1 | 0.000 |
| | | | Above | 0.131 (0.01) (0.11, 0.15) | 292.3 | 14.8 | 0.000 | 0.131 (0.01) (0.11, 0.15) | 289.4 | 14.6 | 0.000 |

Tfh: T follicular helper cell, Th2: T helper 2 cell, Th1: T helper 1 cell, Treg: T regulatory cell, Th17: T helper 17 cell, PFOS: perfluorooctanesulfonic acid, PFOA: perfluorooctanoic acid, PFNA: perfluorononanoic acid, PFHxS: perfluorohexane sulfonic acid, LSMEAN: least-square means, SE: standard error, CI: confidence intervals, df: degrees of freedom. PFAS Value: below and above the PFAS median value of the population.

**Supplemental Table 10:** Unadjusted and adjusted pairwise comparison of the predicted marginal mean between categorical maternal PFAS for differences-over-time models.

| Cell Type | PFAS | Contrast | Visit | Unadjusted | | | | Adjusted | | | |
|---|---|---|---|---|---|---|---|---|---|---|---|
| | | | | Estimate (SE) | df | t-ratio | P-value | Estimate (SE) | df | t-ratio | P-value |
| Tfh | PFOS | Median PFAS (Below) – (Above) | Birth | 0.001 (0.05) | 275.3 | 0.0 | 0.977 | -0.030 (0.05) | 265.4 | -0.6 | 0.522 |
| | | | Month-6 | 0.063 (0.07) | 277.2 | 0.9 | 0.364 | 0.039 (0.07) | 280.2 | 0.6 | 0.582 |
| | | | Month-12 | 0.269 (0.08) | 259.6 | 3.5 | 0.001 | 0.242 (0.08) | 261.4 | 3.1 | 0.002 |
| | PFOA | Median PFAS (Below) – (Above) | Birth | 0.050 (0.05) | 274.3 | 1.1 | 0.281 | 0.034 (0.05) | 256.5 | 0.7 | 0.500 |
| | | | Month-6 | 0.041 (0.07) | 278.5 | 0.6 | 0.555 | 0.015 (0.07) | 285.7 | 0.2 | 0.832 |
| | | | Month-12 | 0.102 (0.08) | 259.2 | 1.3 | 0.195 | 0.086 (0.08) | 268.4 | 1.1 | 0.293 |
| | PFNA | Median PFAS (Below) – (Above) | Birth | -0.005 (0.05) | 274.5 | -0.1 | 0.910 | -0.027 (0.05) | 268.4 | -0.6 | 0.565 |
| | | | Month-6 | -0.010 (0.07) | 278.1 | -0.1 | 0.881 | -0.033 (0.07) | 277.2 | -0.5 | 0.643 |
| | | | Month-12 | 0.156 (0.08) | 260.8 | 2.0 | 0.045 | 0.142 (0.08) | 263.4 | 1.8 | 0.069 |
| | PFHXS | Median PFAS (Below) – (Above) | Birth | -0.008 (0.05) | 274.4 | -0.2 | 0.866 | -0.016 (0.05) | 269.4 | -0.3 | 0.734 |
| | | | Month-6 | 0.013 (0.07) | 278.1 | 0.2 | 0.850 | -0.003 (0.07) | 277.8 | 0.0 | 0.969 |
| | | | Month-12 | 0.049 (0.08) | 259.5 | 0.6 | 0.534 | 0.029 (0.08) | 260.6 | 0.4 | 0.712 |
| Th2 | PFOS | Median PFAS (Below) – (Above) | Birth | 0.002 (0.03) | 291.1 | 0.1 | 0.954 | -0.022 (0.03) | 282.6 | -0.7 | 0.467 |
| | | | Month-6 | -0.047 (0.05) | 289.8 | -1.0 | 0.300 | -0.074 (0.05) | 289.4 | -1.6 | 0.107 |
| | | | Month-12 | -0.098 (0.05) | 284.1 | -1.9 | 0.055 | -0.133 (0.05) | 283.0 | -2.6 | 0.011 |
| | PFOA | Median PFAS (Below) – (Above) | Birth | 0.004 (0.03) | 291.2 | 0.1 | 0.893 | -0.032 (0.03) | 277.1 | -1.1 | 0.290 |
| | | | Month-6 | -0.076 (0.04) | 290.5 | -1.7 | 0.087 | -0.118 (0.05) | 290.9 | -2.6 | 0.010 |
| | | | Month-12 | -0.118 (0.05) | 284.7 | -2.3 | 0.020 | -0.164 (0.05) | 287.3 | -3.1 | 0.002 |
| | PFNA | Median PFAS (Below) – (Above) | Birth | 0.003 (0.03) | 291.1 | 0.1 | 0.904 | -0.012 (0.03) | 285.1 | -0.4 | 0.671 |
| | | | Month-6 | -0.076 (0.04) | 290.2 | -1.7 | 0.090 | -0.100 (0.05) | 287.9 | -2.2 | 0.028 |
| | | | Month-12 | -0.154 (0.05) | 284.9 | -3.1 | 0.002 | -0.170 (0.05) | 284.3 | -3.4 | 0.001 |
| | PFHXS | Median PFAS (Below) – (Above) | Birth | 0.001 (0.03) | 291.2 | 0.0 | 0.961 | -0.008 (0.03) | 285.9 | -0.3 | 0.784 |
| | | | Month-6 | -0.041 (0.05) | 290.4 | -0.9 | 0.367 | -0.050 (0.05) | 288.1 | -1.1 | 0.274 |
| | | | Month-12 | -0.064 (0.05) | 284.8 | -1.3 | 0.210 | -0.080 (0.05) | 283.7 | -1.5 | 0.123 |
| Th1 | PFOS | Median PFAS (Below) – (Above) | Birth | 0.007 (0.02) | 291.4 | 0.4 | 0.684 | 0.006 (0.02) | 281.5 | 0.3 | 0.742 |
| | | | Month-6 | 0.029 (0.03) | 290.2 | 1.1 | 0.291 | 0.028 (0.03) | 288.8 | 1.0 | 0.307 |
| | | | Month-12 | -0.027 (0.03) | 284.9 | -0.9 | 0.373 | -0.027 (0.03) | 281.3 | -0.9 | 0.386 |
| | PFOA | Median PFAS (Below) – (Above) | Birth | 0.010 (0.02) | 291.3 | 0.6 | 0.572 | 0.011 (0.02) | 274.8 | 0.6 | 0.562 |
| | | | Month-6 | 0.010 (0.03) | 290.6 | 0.4 | 0.710 | 0.008 (0.03) | 290.6 | 0.3 | 0.761 |
| | | | Month-12 | -0.040 (0.03) | 284.9 | -1.3 | 0.183 | -0.039 (0.03) | 285.1 | -1.2 | 0.222 |

| | | | | | | | | | | | |
|---|---|---|---|---|---|---|---|---|---|---|---|
| | PFNA | Median PFAS (Below) – (Above) | Birth | 0.010 (0.02) | 291.4 | 0.6 | 0.549 | 0.010 (0.02) | 284.2 | 0.6 | 0.572 |
| | | | Month-6 | 0.002 (0.03) | 290.6 | 0.1 | 0.937 | 0.002 (0.03) | 287.2 | 0.1 | 0.955 |
| | | | Month-12 | -0.071 (0.03) | 285.6 | -2.4 | 0.019 | -0.072 (0.03) | 282.8 | -2.4 | 0.019 |
| | PFHXS | Median PFAS (Below) – (Above) | Birth | 0.003 (0.02) | 291.2 | 0.2 | 0.877 | 0.004 (0.02) | 284.9 | 0.2 | 0.808 |
| | | | Month-6 | 0.016 (0.03) | 290.4 | 0.6 | 0.548 | 0.015 (0.03) | 287.3 | 0.6 | 0.580 |
| | | | Month-12 | -0.042 (0.03) | 284.7 | -1.4 | 0.172 | -0.043 (0.03) | 281.9 | -1.4 | 0.164 |
| Treg | PFOS | Median PFAS (Below) – (Above) | Birth | -0.008 (0.02) | 293.9 | -0.3 | 0.740 | -0.008 (0.02) | 285.5 | -0.3 | 0.730 |
| | | | Month-6 | 0.036 (0.04) | 293.4 | 1.0 | 0.323 | 0.036 (0.04) | 290.7 | 1.0 | 0.336 |
| | | | Month-12 | -0.057 (0.04) | 291.5 | -1.4 | 0.162 | -0.057 (0.04) | 287.4 | -1.4 | 0.178 |
| | PFOA | Median PFAS (Below) – (Above) | Birth | 0.012 (0.02) | 293.8 | 0.5 | 0.596 | 0.013 (0.02) | 279.8 | 0.5 | 0.591 |
| | | | Month-6 | -0.019 (0.04) | 293.5 | -0.5 | 0.603 | -0.020 (0.04) | 291.0 | -0.5 | 0.600 |
| | | | Month-12 | -0.043 (0.04) | 291.4 | -1.1 | 0.293 | -0.041 (0.04) | 289.5 | -1.0 | 0.339 |
| | PFNA | Median PFAS (Below) – (Above) | Birth | 0.013 (0.02) | 293.6 | 0.6 | 0.571 | 0.013 (0.02) | 287.3 | 0.6 | 0.568 |
| | | | Month-6 | -0.013 (0.04) | 293.2 | -0.4 | 0.722 | -0.013 (0.04) | 289.5 | -0.4 | 0.721 |
| | | | Month-12 | -0.109 (0.04) | 291.0 | -2.7 | 0.007 | -0.111 (0.04) | 287.7 | -2.7 | 0.007 |
| | PFHXS | Median PFAS (Below) – (Above) | Birth | -0.014 (0.02) | 294.0 | -0.6 | 0.522 | -0.013 (0.02) | 288.7 | -0.6 | 0.556 |
| | | | Month-6 | -0.068 (0.04) | 293.7 | -1.9 | 0.057 | -0.069 (0.04) | 290.2 | -1.9 | 0.054 |
| | | | Month-12 | -0.127 (0.04) | 292.1 | -3.2 | 0.002 | -0.130 (0.04) | 288.8 | -3.2 | 0.002 |
| Th17 | PFOS | Median PFAS (Below) – (Above) | Birth | 0.000 (0.01) | 294.2 | 0.0 | 0.970 | -0.001 (0.01) | 286.5 | -0.1 | 0.881 |
| | | | Month-6 | 0.002 (0.01) | 293.8 | 0.2 | 0.852 | 0.000 (0.01) | 290.9 | 0.0 | 0.973 |
| | | | Month-12 | 0.002 (0.01) | 292.3 | 0.1 | 0.886 | -0.001 (0.01) | 288.9 | -0.1 | 0.948 |
| | PFOA | Median PFAS (Below) – (Above) | Birth | 0.000 (0.01) | 294.3 | 0.0 | 0.999 | -0.001 (0.01) | 281.7 | -0.2 | 0.862 |
| | | | Month-6 | 0.008 (0.01) | 294.1 | 0.7 | 0.505 | 0.005 (0.01) | 290.7 | 0.4 | 0.665 |
| | | | Month-12 | 0.021 (0.01) | 292.8 | 1.6 | 0.111 | 0.020 (0.01) | 290.6 | 1.4 | 0.152 |
| | PFNA | Median PFAS (Below) – (Above) | Birth | 0.001 (0.01) | 294.1 | 0.1 | 0.940 | 0.000 (0.01) | 288.4 | 0.0 | 0.961 |
| | | | Month-6 | 0.011 (0.01) | 293.8 | 0.9 | 0.352 | 0.009 (0.01) | 290.3 | 0.7 | 0.467 |
| | | | Month-12 | -0.007 (0.01) | 292.3 | -0.5 | 0.595 | -0.009 (0.01) | 289.3 | -0.7 | 0.492 |
| | PFHXS | Median PFAS (Below) – (Above) | Birth | 0.000 (0.01) | 294.0 | 0.0 | 0.984 | 0.000 (0.01) | 289.0 | 0.0 | 0.976 |
| | | | Month-6 | -0.001 (0.01) | 293.7 | -0.1 | 0.904 | -0.002 (0.01) | 290.4 | -0.1 | 0.896 |
| | | | Month-12 | -0.017 (0.01) | 292.0 | -1.3 | 0.205 | -0.019 (0.01) | 289.3 | -1.4 | 0.170 |

Tfh: T follicular helper cell, Th2: T helper 2 cell, Th1: T helper 1 cell, Treg: T regulatory cell, Th17: T helper 17 cell, PFOS: perfluorooctanesulfonic acid, PFOA: perfluorooctanoic acid, PFNA: perfluorononanoic acid, PFHxS: perfluorohexane sulfonic acid, SE: standard error, df: degrees of freedom.

**Supplemental Table 11:** Unadjusted and adjusted interaction P-values between categorical maternal PFAS and infant age for differences-over-time models.

| Cell Type | PFAS | Unadjusted P-value | Adjusted P-value |
|---|---|---|---|
| Tfh | PFOS | 0.004 | 0.003 |
| | PFOA | 0.781 | 0.738 |
| | PFNA | 0.109 | 0.089 |
| | PFHXS | 0.783 | 0.860 |
| Th2 | PFOS | 0.180 | 0.119 |
| | PFOA | 0.052 | 0.031 |
| | PFNA | 0.011 | 0.010 |
| | PFHXS | 0.438 | 0.393 |
| Th1 | PFOS | 0.355 | 0.359 |
| | PFOA | 0.294 | 0.316 |
| | PFNA | 0.047 | 0.045 |
| | PFHXS | 0.294 | 0.279 |
| Treg | PFOS | 0.214 | 0.218 |
| | PFOA | 0.437 | 0.440 |
| | PFNA | 0.024 | 0.023 |
| | PFHXS | 0.033 | 0.028 |
| Th17 | PFOS | 0.987 | 0.993 |
| | PFOA | 0.353 | 0.360 |
| | PFNA | 0.565 | 0.581 |
| | PFHXS | 0.519 | 0.446 |

Tfh: T follicular helper cell, Th2: T helper 2 cell, Th1: T helper 1 cell, Treg: T regulatory cell, Th17: T helper 17 cell, PFOS: perfluorooctanesulfonic acid, PFOA: perfluorooctanoic acid, PFNA: perfluorononanoic acid, PFHxS: perfluorohexane sulfonic acid, df: degrees of freedom.
